# Synthetic Anatomy: Deep Learning Models for Virtual Population Generation–A Review

**DOI:** 10.1101/2025.10.28.25338782

**Authors:** Zherui Zhou, Arezoo Zakeri, Haoran Dou, Yidan Xue, Michael MacRaild, Jinghan Huang, Fengming Lin, Ali Sarrami-Foroushani, Jinming Duan, Alejandro F. Frangi

## Abstract

In-silico trials (ISTs) represent a transformative approach in medical research, leveraging computer modelling and simulation to evaluate products virtually. This systematic review examines state-of-the-art methods for generating virtual populations (VPs), a crucial component of ISTs. The review focuses on deep learning techniques developed over the past decade for building generative geometric models specifically designed for the synthesis of virtual organ populations. Through a comprehensive analysis of recent publications, we identified several key challenges in the field: the generation of complex topological structures, effective utilisation of multimodal data, maintenance of cross-resolution consistency, and the absence of standardised evaluation metrics.

We provide a systematic examination of evaluation frameworks, analysing methods across four key dimensions: fidelity, utility, generalisability, and diversity. Additionally, we present a meta-analysis comparing the performance of methods based on different model architectures across various anatomical structures, offering quantitative insights into their relative strengths and limitations. Our review provides a structured examination of data inputs and outputs, generative approaches, and evaluation metrics, whilst discussing the challenges and potential future directions accordingly.

To the best of our knowledge, this work represents the first systematic review specifically focused on geometrical deep learning-based methods for building anatomical virtual populations, offering insights into current limitations and future research directions in this rapidly evolving field.

## 1 Introduction

### 1.1 Background

In-silico trials (ISTs) represent a transformative paradigm in medical research, employing individualised computer modelling and simulation to test medical devices, drugs, and medical procedures virtually [Viceconti et al., 2016]. This computational approach offers unprecedented opportunities to conduct comprehensive assessments of treatment safety and efficacy across diverse patient phenotypes whilst simultaneously addressing the ethical concerns and resource limitations inherent in traditional clinical trials. As regulatory bodies increasingly embrace model-informed evidence frameworks, ensuring the reliability and credibility of IST findings has become paramount for their successful translation into clinical practice.

A fundamental challenge in establishing the reliability of in-silico trials lies in addressing data scarcity and imbalance. Real clinical datasets are inherently constrained by patient access and participation, demographic distributions, geographical differences, and recruitment biases, creating a risk of systematically biased outcomes when used exclusively for virtual experimentation. Virtual populations (VPs), whilst derived from real patient data, offer the potential to encompass broader anatomical, physiological, and pathological variability than traditionally available cohorts. This enhanced diversity makes VPs essential for data balancing and critical for building robust, trustworthy ISTs that can inform regulatory decision-making and clinical practice across heterogeneous patient populations.

### 1.2 Motivation

The generation of realistic virtual populations has emerged as a cornerstone requirement for the success of ISTs, as every virtual patient and virtual chimaera must demonstrate clinical plausibility when compared to real patient cohorts [Viceconti et al., 2020]. This requirement becomes particularly crucial when generating model-informed evidence for healthcare products targeting specific or small subgroups, where traditional clinical trials may prove impractical due to limited participant availability, ethical constraints, or prohibitive costs [Frangi et al., 2023]. The ability to create representative virtual cohorts enables researchers to explore rare disease scenarios, investigate device performance across extreme anatomical variations, and assess treatment outcomes in populations that would be challenging to recruit in conventional trials.

The advent of generative artificial intelligence has revolutionised medical data augmentation and balancing, offering sophisticated approaches to synthetic data creation that were previously unattainable. However, despite these technological advances, a significant gap persists between the development of generative methodologies and their practical application to VP generation for ISTs. Many studies have focused on improving the technical aspects of generative models without adequately addressing the specific requirements and constraints of virtual population synthesis for regulatory and clinical applications.

As demonstrated by the publication timeline in Fig. 3, there has been a marked increase in VP generation research over recent years, reflecting growing recognition of its importance. However, the current landscape faces several critical challenges that impede progress towards clinically meaningful virtual populations. The ambiguous and inconsistent terminology surrounding VP creation for ISTs has led to confusion within the field, hindering the development of cohesive frameworks and standardised methodologies. This terminological inconsistency complicates cross-study comparisons and impedes the establishment of best practices.

Furthermore, evaluating virtual population generation remains a fundamental and largely unresolved challenge. Existing studies typically provide limited assessment approaches, often emphasising visual appearance or basic statistical fidelity rather than clinically meaningful criteria that would ensure the generated populations are suitable for their intended use [Zamzmi et al., 2025]. The absence of standardised evaluation metrics and systematic validation frameworks makes it difficult to rigorously establish the fidelity, utility, generalisability, and diversity of generated populations. This lack of robust evaluation strategies not only undermines confidence in current approaches but also limits the ability to compare different methodologies and identify optimal solutions for specific applications.

The regulatory acceptance of IST evidence further depends on demonstrating that virtual populations exhibit appropriate biological plausibility, maintain relevant correlations between anatomical and physiological parameters, and capture the essential variability present in real patient cohorts. Without comprehensive evaluation frameworks that address these requirements, the translation of VP generation research into clinically applicable tools remains limited.

Addressing these critical gaps, this review provides a comprehensive examination of the current state of virtual population generation for ISTs. We systematically define VP terminology to establish consistent nomenclature within the field, comprehensively map the deep learning landscape for anatomical generation, critically evaluate existing methodologies and their limitations, and provide informed forecasts for the future development of virtual population synthesis technologies. Through this systematic analysis, we aim to guide researchers towards more clinically relevant, reproducible, and regulatory-compliant approaches to virtual population generation.

### 1.3 Definitions

A comprehensive in-silico trial (IST) workflow begins with patient data access and proceeds through several stages before culminating in specific virtual trials and simulation as shown in Fig. 1. Central to this workflow is the generation of VPs, which represent one of the most significant paradigm shifts in computational medicine and regulatory science.

**Figure 1.**
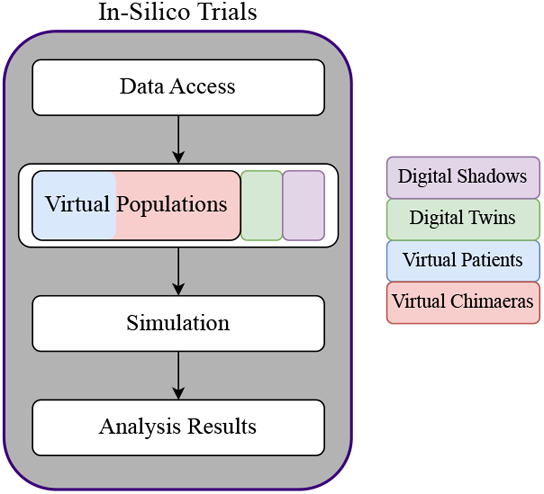
In Silico Trial (IST) workflow and different approaches to build a virtual population

#### Fundamental Concept of Virtual Populations

A VP is derived by instantiating a generative model, *G*(**g**; **m**), to represent a set of digitally synthesised anatomical models in various representations (viz. image, binary map, mesh, point cloud, etc.) modelled from limited real-world population data. These models reflect the inherent variability in anatomical phenotypes (**g**) and associated meta-information (**m**), thereby enabling the systematic exploration of patient diversity that would be impractical or impossible to achieve through traditional clinical studies [Allen et al., 2016].

The mathematical formulation of this generative process captures both the deterministic relationships between anatomical features and the stochastic nature of biological variation. The generative model *G*(**g**; **m**) learns to map from a latent space of anatomical parameters **g** and clinical metadata **m** to realistic patient representations, ensuring that the generated virtual patients maintain physiologically plausible combinations of anatomical structures and pathophysiological states.

#### Taxonomic Classification of Virtual Populations

Virtual populations can be classified through two complementary taxonomic frameworks, each addressing different aspects of their construction and application, as illustrated in Fig. 2.

**Figure 2.**
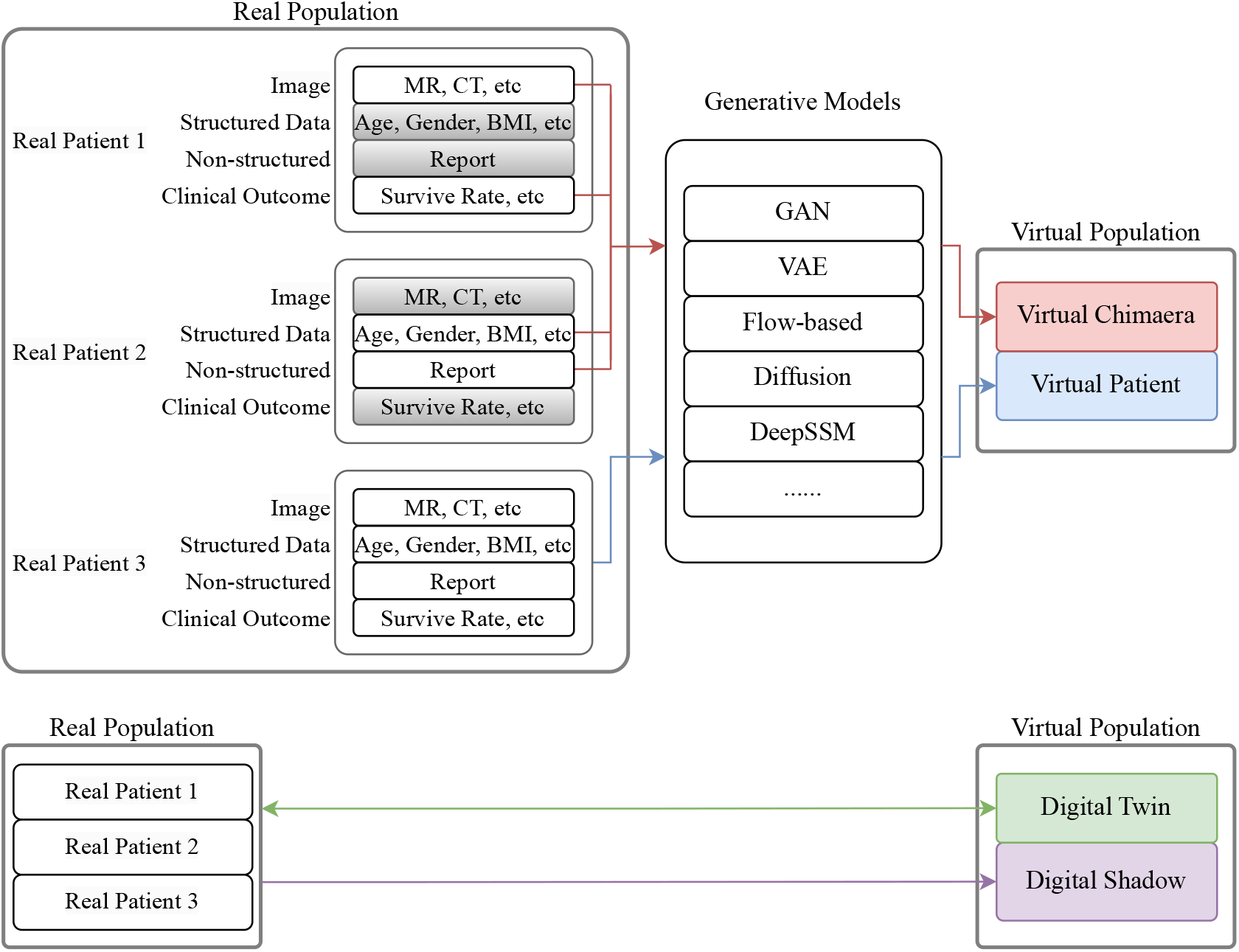
Relationships of Virtual Patients, Virtual Chimaeras, Digital Twins, Digital Shadows, and Virtual Populations. The dark shadow in the Real Population indicates unavailable data.

#### Classification by Data Construction Methodology

The first classification distinguishes between Virtual Patients (VPs) and Virtual Chimaeras (VCs) based on the composition and structure of their training datasets:

- *Virtual Patients* (VPs) represent the traditional approach where the training set corresponds to a single, coherent sample from a real-world population. In this framework, the generative model *G*(**g**; **m**) is trained on data where both anatomical features **g** and metadata **m** originate from the same population cohort [Sinisi et al., 2020]. This approach ensures internal consistency within the generated virtual patients but may be limited by the scope and diversity of the source population.
- *Virtual Chimaeras* (VCs), conversely, represent a more sophisticated approach that addresses the inherent limitations of single-population datasets [Dou et al., 2025]. In this framework, the training set comprises the union of two or more distinct datasets: **g** = (**g**_1_; **g**_2_) and **m** = (**m**_1_; **m**_2_), where traditionally (**g**_1_ ∩ **g**_2_) = ∅ and (**m**_1_ ∩ **m**_2_) = ∅. Each constituent dataset represents a different perspective or “vista” of the same hypothetical population, capturing distinct variables or measurement modalities that collectively encompass the full scope of anatomical and clinical variation the generative model aims to represent.

VCs can also accommodate more complex data fusion scenarios, where the constituent datasets may exhibit partial overlap in their measured variables: (**g**_1_ ∩ **g**_2_) ≠ ∅ and (**m**_1_ ∩ **m**_2_) ≠ ∅ [Dou et al., 2025]. This advancement enables the integration of complementary datasets with shared anatomical landmarks or clinical parameters, thereby enhancing the robustness and generalisability of the resulting virtual population.

#### Classification by Bidirectional Data Exchange Capability

The second classification framework distinguishes between Digital Twins (DTs) and Digital Shadows (DSs) based on the directionality and nature of data exchange between the virtual and physical domains:

- *Digital Twins* feature bidirectional data exchange, creating virtual patient models that both mirror real-time physiological processes and directly inform clinical decision-making [Barricelli et al., 2019]. This continuous synchronisation enables dynamic model updates while allowing virtual insights to guide treatment planning for the corresponding physical patient.
- *Digital Shadows* maintain high-fidelity patient representations but with unidirectional data flow from physical to digital domains only [Sepasgozar, 2021]. These passive models support retrospective analysis and population studies without directly influencing patient treatment or monitoring.

#### Synthesis and Clinical Implications

The intersection of these classification frameworks creates a comprehensive taxonomy that enables researchers to select the most appropriate virtual population methodology for their specific applications. The choice between Virtual Patients and Virtual Chimaeras depends primarily on data availability and the desired scope of population representation, whilst the selection between Digital Twins and Digital Shadows is determined by the intended clinical workflow and the requirement for real-time bidirectional interaction.

### 1.4 Inclusion and Exclusion Criteria

Virtual population construction typically involves selecting relevant data inputs, learning generative models, and instantiating them with conditioning variables to synthesise diverse virtual patients. The following criteria were applied to identify relevant publications.

#### Inclusion Criteria

a) Studies employing generative models for shape synthesis; b) Population-level anatomical data generation; c) Deep learning-based 3-dimensional (3D) generation approaches; d) Human medical applications; e) Publications from 2015-2025.

### Exclusion Criteria

a) Animal/veterinary studies; b) Pure image generation without geometric modelling; c) Reconstruction-only tasks lacking generative components.

### 1.5 Review Methodology

This systematic review followed a protocol registered on PROSPERO (CRD42024605863). The initial search yielded 3611 records, subsequently screened according to PRISMA guidelines^1^, resulting in 51 studies meeting all inclusion criteria as shown in Fig. 4. We briefly introduce generative models based on deep learning and their variants in Section 2, provide specific methods applied to anatomical shapes following our taxonomies in Section 3 (Fig. 5), and introduce evaluation metrics in Section 4. Section 5 presents a meta-analysis of collected studies, Section 6 discusses challenges and potential future directions based on our analysis, and Section 7 concludes this review.

**Figure 3.**
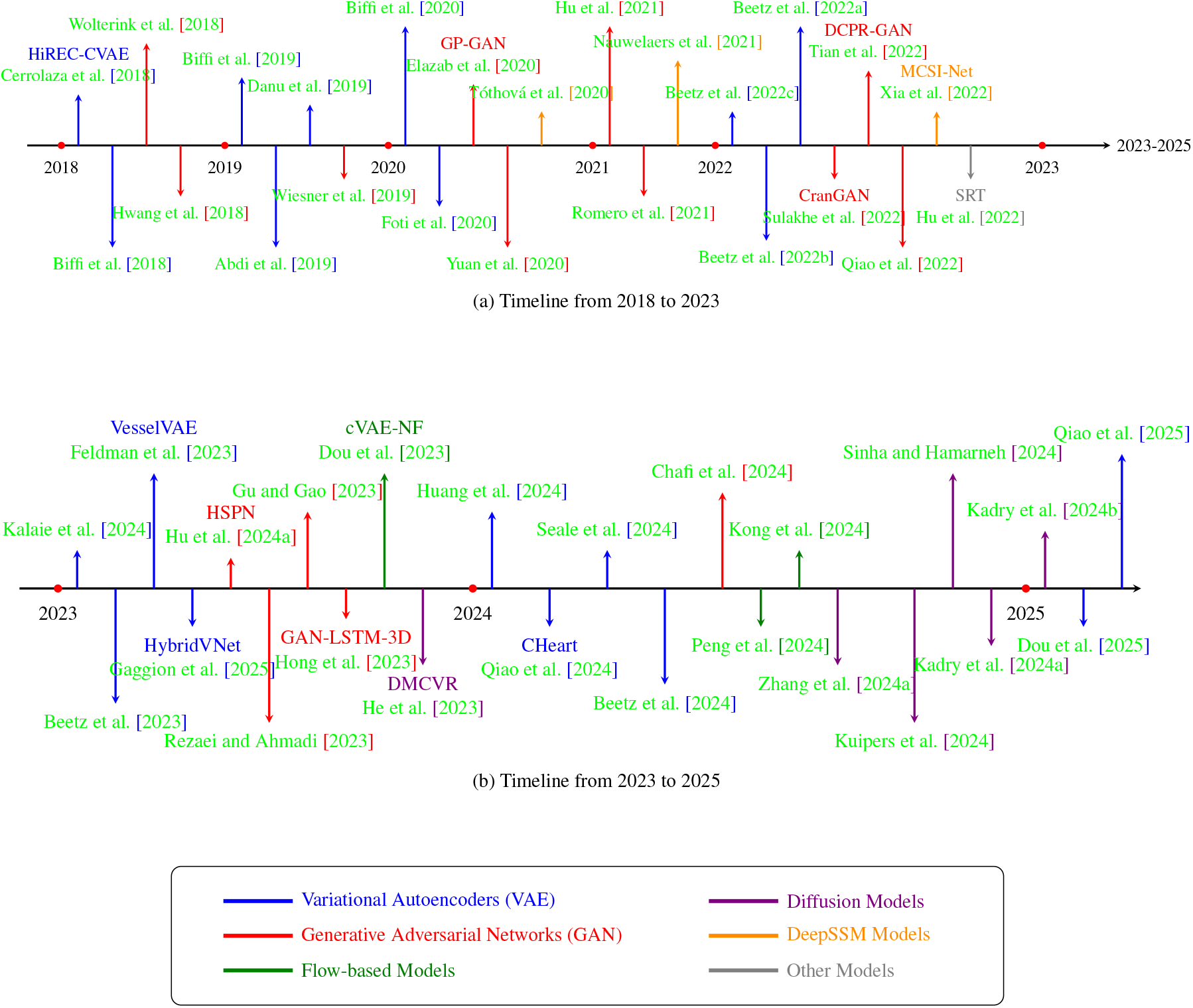
Timeline of deep learning-based generative models for virtual population generation (2018-2025). No publications that match the inclusion and exclusion criteria were found earlier than 2018.

**Figure 4.**
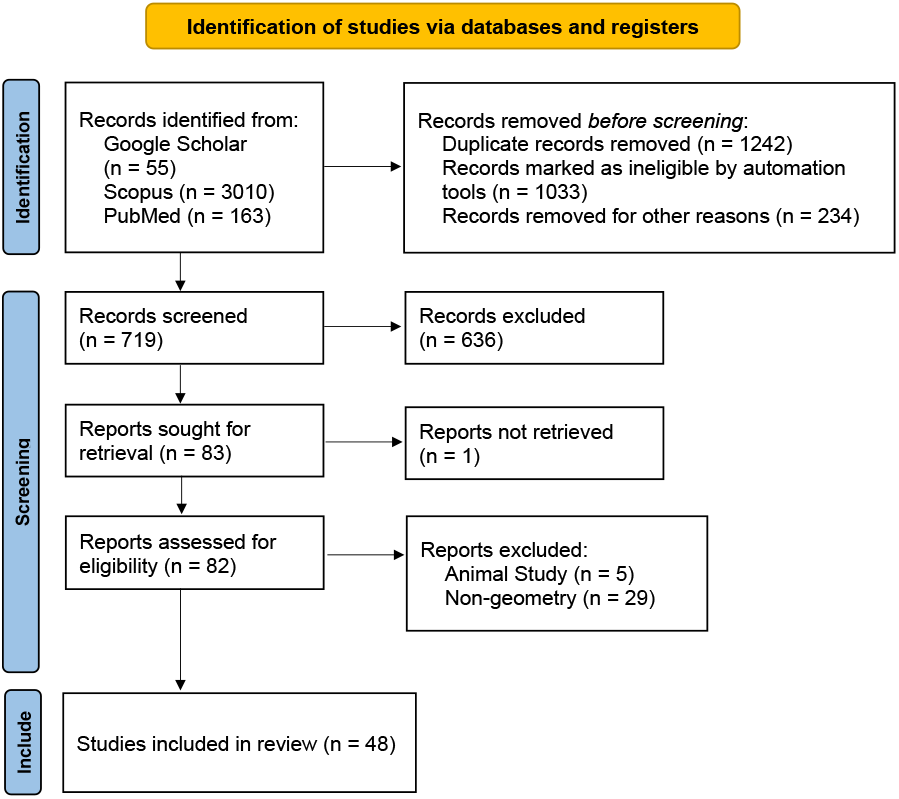
Prisma Flow Diagram

**Figure 5.**
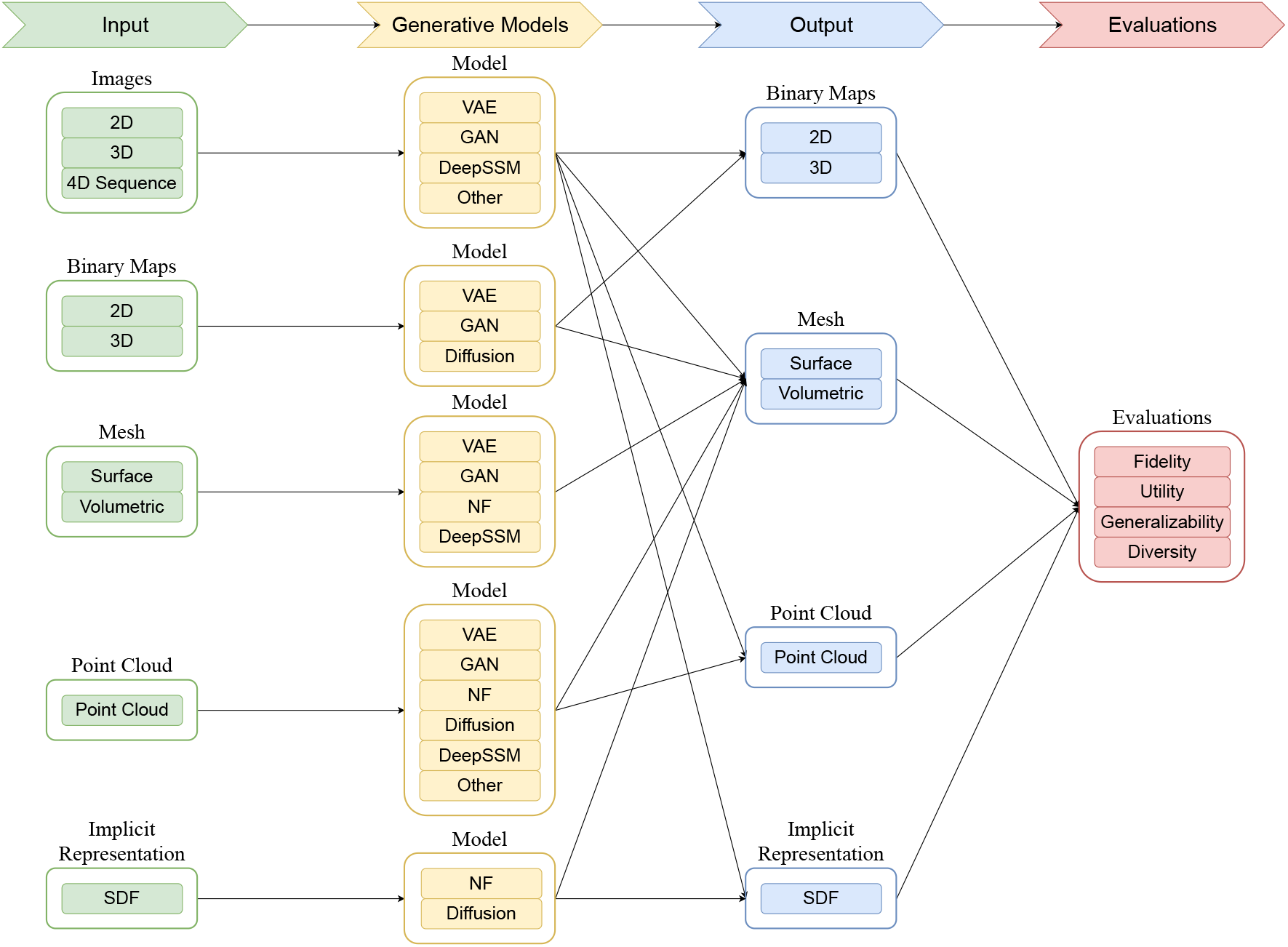
Taxonomy of Deep Learning-Based Generative Methods

### 1.6 Contributions

Previous reviews have primarily focused on generative models, medical imaging, or the evaluation of synthetic data. For example, both Xiao et al. [2020] and Xu et al. [2023] summarised advances in 3D deep geometry learning, with the former emphasising representation approaches and the latter emphasising model development; however, neither specifically addressed anatomical geometries. Singh and Raza [2021] reviewed the use of Generative Adversarial Networks (GANs) in medical imaging, but the scope was limited to GAN-based methods and image representations. Bandi et al. [2023] provided a comprehensive overview of generative models, yet a comparable synthesis is still lacking in the context of deep learning for anatomical geometry generation. Reviews such as Figueira and Vaz [2022], Bandi et al. [2023], Osorio-Marulanda et al. [2024] discussed the evaluation of synthetic data, but not from a clinical standpoint. More recently, Liu et al. [2024] bridged the gap between 3D generative modelling and medical imaging by introducing generative applications to 3D brain and heart volumetric data; however, the models discussed therein extend beyond generation to include tasks such as segmentation and registration. Lin et al. [2025] surveyed image-to-mesh reconstruction methods for anatomical geometry, which overlaps with our review in application scope; however, their survey does not address generative modelling. To the best of our knowledge, this work represents the first systematic review of deep generative models of anatomy and their evaluation. Our main contributions are summarised as follows:

- We present a comprehensive list of definitions for VP terminologies.
- We introduce fundamental models and their variants applicable to virtual patient population generation, providing a classification of state-of-the-art methods based on these foundational models.
- We compile and categorise virtual population evaluation metrics according to our systematic evaluation standards of fidelity, utility, generalizability, and diversity (please refer to Section IV for the definitions of these terms). Additionally, we present a meta-analysis comparing different metrics across anatomies and model bases.
- We identify current challenges and future directions of VP generation.

## 2 Traditional Statistical Shape Models

Although this paper focuses on deep-learning methods, we will still briefly introduce the traditional statistical shape models, their application to VPs, and their limitations.

Statistical Shape Models (SSMs) provide a mathematical and data-driven framework for capturing and analysing the variability of anatomical structures across populations [Ambellan et al., 2019]. Constructed from 3D reconstructions of medical imaging data, SSMs represent both the mean anatomical shape and its principal modes of variation. By establishing point-wise correspondences across samples, SSMs allow the systematic study of normal and pathological morphological variability, growth, and disease progression [Adams et al., 2023].

They have been applied to diverse anatomical structures—including hearts [Mitchell et al., 2001, Lötjönen et al., 2004, Ordas et al., 2007, Grbic et al., 2012], livers [Lamecker et al., 2002, Okada et al., 2008], joints [Pedoia et al., 2015, Baldwin et al., 2010, van Buuren et al., 2021, Balestra et al., 2014], and brains [Styner et al., 2006, Tao et al., 2002, Shen et al., 2001] to support geometry generations, population-level shape analysis, and longitudinal studies of anatomical change. These models enable applications ranging from computer-assisted diagnosis, surgical planning, implant design, and therapy evaluation to medical education and the creation of digital anatomy atlases.

Still, their performance is often limited when it comes to shapes with complicated details and topologies like blood vessel trees [Du, 2018]. This is because the principal component analysis (PCA) that relies on shape variation lies in a linear subspace. Thus, deep-learning methods are required to deal with non-linear, multi-scale, or appearance-driven variability.

## 3 Deep Learning Based Generative Models

In comparison with traditional methods, deep learning based generative models have significantly advanced geometry representation and virtual population modelling by enabling the creation of complex, realistic shapes and anatomical structures. These models vary widely in their architectures, training paradigms, and target applications, making them versatile tools for geometry generation.

The architectural diversity of these models dictates their functionality and suitability for specific tasks. Variational autoencoders (VAEs) [Kingma and Welling, 2022] rely on probabilistic modelling and an encoder-decoder structure to learn latent representations. Generative adversarial networks (GANs) [Goodfellow et al., 2014] use an adversarial training framework involving two neural networks: a generator that tries to produce realistic samples, and a discriminator that attempts to distinguish between real samples from the training data and fake samples produced by the generator. Diffusion models [Ho et al., 2020] leverage iterative noise addition and denoising processes to generate new shapes. Normalising flow models [Rezende and Mohamed, 2015] employ invertible transformations to map between simple and complex distributions. Deep Learning-based Statistical Shape Models (DeepSSMs) integrate traditional statistical techniques with deep learning networks, offering interpretable and parameter-efficient approaches to model shape variability. In the rest of this section, we will introduce each fundamental model and its variants in detail.

### 3.1 Variational AutoEncoder

VAE is a type of probabilistic generative model that learns latent representations of data using an encoder-decoder framework. Unlike traditional autoencoders, VAEs learn a continuous latent space that enables both meaningful interpolation and the generation of new geometric data. The inherent probabilistic nature of this framework makes it particularly suitable for medical geometry generation, where understanding uncertainty and capturing anatomical features and variations are crucial.

The architecture consists of two primary components: an encoder network and a decoder network. As shown in Fig. 6, the encoder maps the ground truth geometry *x* to a latent distribution *q*_*ϕ*_(*z*|*x*), effectively approximating the true posterior *p*_*θ*_(*z*|*x*). It outputs the mean *µ* and variance *σ*^2^ of the latent Gaussian distribution, enabling inference through a reparameterization technique. The decoder network, serving as the generative model, maps latent features *z* sampled from the learned distribution to generate VP *x*^*′*^, modelling the likelihood *p*_*θ*_(*x*|*z*).

**Figure 6.**
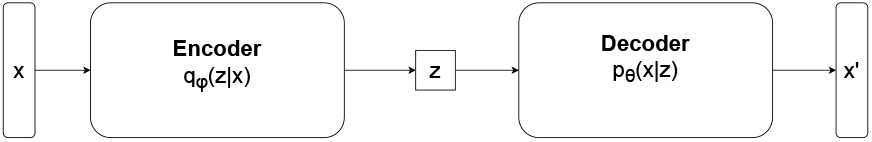
VAE Structure

The VAE optimisation objective combines reconstruction accuracy with distribution matching. The loss function consists of two terms, a reconstruction loss ensuring geometric fidelity, and a Kullback-Leibler (KL) divergence term regularising the learned latent distribution towards the prior, which is formulated as:

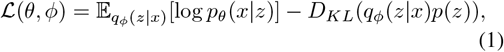

where *θ* represents the parameters of the decoder network, *ϕ* represents the parameters of the encoder network, *x* is the input data, *z* is the latent vector, *q*_*ϕ*_(*z*|*x*) is the encoder’s approximate posterior distribution, *p*_*θ*_(*x*|*z*) is the decoder’s likelihood distribution, *p*(*z*) is the prior distribution, typically a standard normal distribution 𝒩 (0, *I*), *D*_*KL*_ is the KL divergence between the approximated posterior and prior distributions of latent variables. To enable back-propagation through the sampling process, VAEs employ the reparameterization trick:

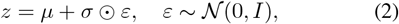

where *z* is the sampled latent vector, *µ* is the mean vector output by the encoder, *σ* is the standard deviation vector output by the encoder, ⊙ represents element-wise multiplication, *ε* is a random noise vector sampled from a standard normal distribution, 𝒩 (0, *I*) denotes a multivariate normal distribution with zero mean and identity covariance matrix.

The basic VAE architecture has been extended in several ways to better handle geometric data and address specific limitations, such as conditional VAE (cVAE) [Sohn et al., 2015] which improves the standard VAE by incorporating additional conditioning information into both encoding and decoding processes.

Recurrent variational autoencoder (RVAE) [Chung et al., 2015], combines VAE with recurrent neural networks to handle sequential or temporal geometric data.

Mesh VAE [Litany et al., 2018], is specially designed for mesh representations. It uses Graph Convolutional Networks as encoder and decoder, allowing VAE to explicitly preserve mesh topology and directly generate vertex positions and connectivity.

Ladder VAE (LVAE) [Sønderby et al., 2016] introduces a hierarchical structure to the latent space, where different levels of the hierarchy capture different levels of abstraction in the data.

### 3.2 Generative Adversarial Networks

Generative Adversarial Networks embody a revolutionary approach to geometric data generation through an adversarial learning framework. At its core, GANs consist of two neural networks engaged in a competitive learning process, structured as a two-player minimax game as shown in Fig. 7, where the ultimate goal is to reach a Nash equilibrium between the competing networks.

**Figure 7.**
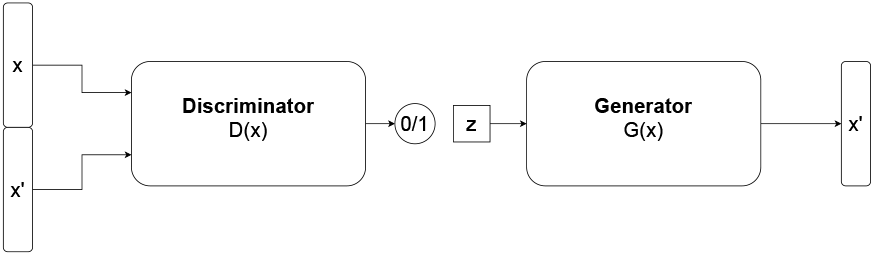
GAN Structure

**Figure 8.**
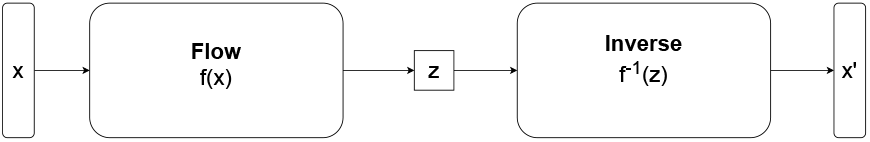
Normalising Flow Structure

**Figure 9.**
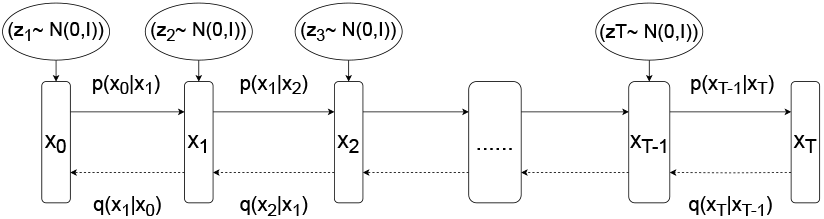
Diffusion Model Structure

The Generator (*G*) serves as the generative model within the GAN framework. The goal of the GAN is to closely resemble the probability distribution of the training samples and to generate new samples using a random noise vector drawn from a prior Gaussian or uniform distribution. The noise serves as the seed from which synthetic geometric data emerges. The Discriminator (*D*) acts as the critical evaluator, learning to distinguish between real geometric data and the generator’s synthetic creations. After receiving input data, the *D* outputs a probability representing the likelihood that the input data originates from the real distribution rather than being synthetically generated. During the training phase, the *D* learns to assign high probabilities to real data and low probabilities to generated samples, and *G* is trained to create samples that the discriminator will mistake for real data. The training process can be mathematically expressed through the value function:

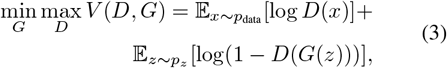

where *x* is the real population samples from the training dataset, *z* is the random noise vector input to *G, p*_data_ is the real data distribution, *p*_*z*_ is the prior noise distribution. *x* ∼ *p*_data_ indicates *x* is sampled from the real data distribution, and *z* ∼ *p*_*z*_ indicates *z* is sampled from the noise distribution. min_*G*_ max_*D*_ suggests that G minimises while D maximises the objective, and the value function *V* (*D, G*) defines the competition between D and G.

Several variants of GANs are proposed to solve the challenges that standard GANs are facing. Conditional GAN (cGAN) [Mirza and Osindero, 2014] extends the GAN framework by incorporating additional conditioning information 𝒸 into both the generator and discriminator networks.

Wasserstein GAN (WGAN) [Arjovsky et al., 2017] addresses fundamental stability issues in GAN training by adopting the Wasserstein distance as a metric.

Sphere-guided GAN (SP-GAN) [Li et al., 2021a] is specifically designed for 3D shape generation and manipulation, using sphere guidance as its core innovation.

CycleGAN [Zhu et al., 2017] is a framework originally for unpaired image-to-image translation that learns to map between two domains without paired training data; it can also be implemented for VP generation.

### 3.3 Normalising Flows

Normalising flows represent a class of generative models that learn invertible transformations between a simple base distribution (Input) and a complex target distribution. Unlike GANs and VAEs, the encoder and decoder in normalising flows are the exact inverse versions of each other, as shown in 8, making them suitable for geometric VP generation where bidirectional transformation is desired [Rezende and Mohamed, 2015, Dinh et al., 2017]. The core principle of normalising flows is based on the change of variables formula:

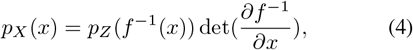

where *p*_*X*_ (*x*) is the target distribution we want to model, *p*_*Z*_ is a simple base distribution (typically Gaussian), *f* is an invertible transformation, and the Jacobian determinant term det (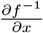) accounts for the volume change caused by the invertible transformation. This determinant term plays a crucial role in normalising flows by ensuring the transformed probability distribution remains properly normalised. This forms the basis of the training objective, where we maximise the log-likelihood of the observed data. The negative log-likelihood loss for a batch of N samples is:

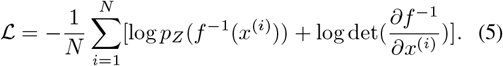

When generating, *z* will be sampled from *p*_*Z*_ and transformed to *x* = *f* (*z*). A technique closely related to normalising flows, especially to continuous normalising flow (CNF), is neural ordinary differential equations (NODEs) [Chen et al., 2018]. In CNF, NODEs extend the idea of normalising flows into a continuous domain. This approach parameterises the continuous dynamics of hidden states using neural networks, rather than applying discrete transformations through traditional layers. The core concept of Neural ODEs is to define the evolution of hidden states through a differential equation:

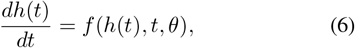

where *f* is a neural network that computes the derivative of the hidden state *h*(*t*) with respect to time *t*, and *θ* represents the network parameters. The transformation from input to output is then obtained by solving this ODE from an initial time *t*_0_ to a final time *t*_1_:

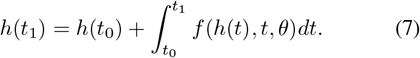

This enables the model to handle irregular time series data and helps process medical image sequences.

### 3.4 Diffusion Models

Diffusion Models represent a class of generative models that learn to gradually denoise data through an iterative process. The model operates by progressively corrupting training data through the addition of Gaussian noise, and then learning to reverse this noise corruption process [Bandi et al., 2023], as shown in 9. Once learned, the model is able to denoise a randomly sampled Gaussian noise to generate valid samples that match the training data distribution [Zhang et al., 2023a]. For geometry generation, this approach offers high-quality results with stable training dynamics. The core principle of diffusion models involves two processes: a forward diffusion process that gradually adds noise to the data, and a reverse process that learns to denoise it. The forward process defines a sequence of progressively noisier versions of the data, while the reverse process learns to recover the original data structure step by step. The forward diffusion process is formulated as a Markov chain that gradually adds Gaussian noise to the data:

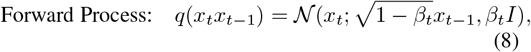

where *x*_*t*_ represents the data at timestep *t*, and *β*_*t*_ is a carefully chosen noise schedule that controls the rate of noise addition. This process can be applied for *T* steps, eventually transforming any data distribution into approximately Gaussian noise. The reverse process, which is learned during training, attempts to reverse this noise corruption:

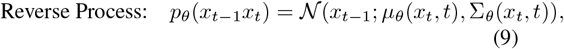

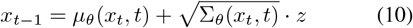

where *µ*_*θ*_ and Σ_*θ*_ are neural networks that predict the parameters of the reverse process, *µ*_*θ*_(*x*_*t*_, *t*) and Σ_*θ*_(*x*_*t*_, *t*) are the predicted mean and variance, and *z* is random Gaussian noise. The model learns to estimate the noise that was added at each step, effectively learning how to gradually denoise the data. While the original training objective is derived from a variational lower bound, in practice, it is simplified to a remarkably effective objective:

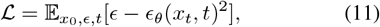

in which the model is trained to predict the noise *ϵ* that was added to the data at each timestep.

Original diffusion models face slow sampling speed and optimisation difficulties. The reverse diffusion process requires hundreds to thousands of iterative steps to refine noisy data into realistic samples. Each step involves running a large neural network, making the process computationally slow, and the loss optimisation is harder to perform when the reverse process is closer to the data distribution. Denoising Diffusion Probabilistic Models (DDPM) introducing a fixed variance schedule that follows either a linear or cosine function.

Score-based Generative Models (SGMs) reformulate diffusion models in terms of score matching, focusing on learning the gradient of the log density.

Another variant is Latent Diffusion Models (LDMs), which operate in a compressed latent space. It learns the data distribution in a more computationally efficient way by applying the diffusion process to the latent space.

Denoising Diffusion Implicit Models (DDIMs) [Song et al., 2022] modify the traditional diffusion process by introducing a non-Markovian inference procedure that enables faster sampling while maintaining generation quality.

### 3.5 Deep Learning Based Statistical Shape Models

Statistical Shape Models (SSMs) represent a foundational approach in geometric modelling that captures shape variations within a population through statistical analysis. Traditional SSMs typically employ PCA to model shape variations in a linear subspace, where shapes are represented as points in a high-dimensional space defined by corresponding landmarks, as in point distribution models (PDMs). The integration of deep learning with SSMs has addressed several limitations of conventional approaches, particularly their linear nature and requirement for precise correspondences. Deep Learning-Based Statistical Shape Models (DeepSSMs) leverage neural networks’ capacity to learn complex, non-linear shape variations while maintaining the interpretable nature of traditional SSMs [Bhalodia et al., 2018]. A basic structure of the DeepSSMs is as Fig. 10. It can be formulated as:

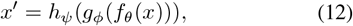

where *f*_*θ*_(*x*) is the encoder (could be CNN, GCN, etc.) with parameters *θ* to learn the shape parameters of the real population, *g*_*ϕ*_ is a transformation function that transforms the latent representation into statistical shape parameters, and *h*_*ψ*_ is the SSM to generate VP from the shape parameters.

**Figure 10.**
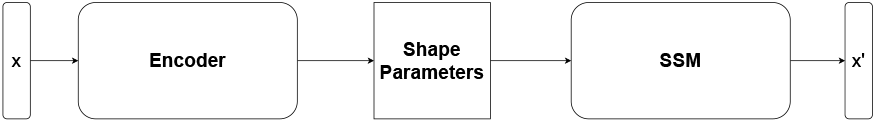
Deep Learning SSM Structure

These models typically use architectures like autoen-coders, CNN, GCN, or other deep learning models to learn latent representations that encode shape variability. CNN and GCN have been applied to the medical area in regression tasks like segmentation [Kayalibay et al., 2017], classification [Li et al., 2014], detection [Tiwari et al., 2022], and prediction, proving their abilities to capture complex features. Combined with SSMs (Point Distribution Models, Statistical Deformation Models, etc.), DeepSSMs ensure interpretability while leveraging the representational power of deep learning.

## 4 Generative Methods Classification

In recent years, deep learning-based methods have emerged as powerful tools for modelling and generating anatomical geometries, which in this review are mesh, point cloud, 3D binary map, and implicit representations. This section presents a comprehensive classification of these methods, encompassing both direct generation approaches for virtual populations and related applications that, while not explicitly focused on population generation, offer valuable insights and transferable techniques. We organised our review primarily along two fundamental axes: the nature of the generation process (unconditional versus conditional) and the underlying deep learning architectures employed, which will be classified as previously in Section 2: VAEs, GANs, Flow-based models, Diffusion models, and DeepSSMs. This organisation is particularly relevant because conditional generation is necessary for in-silico trials, as virtual population generation is often applied to augment imbalanced data, which means control over the generation process is required to produce synthetic data with specific features. For each category, we first examine methods specifically designed for generation tasks, followed by a discussion of relevant non-generation applications using generative models that demonstrate promising techniques adaptable to virtual population synthesis. While some generative models were specifically developed for virtual population synthesis, others were initially designed for related tasks such as shape completion, reconstruction, or prediction. We include these adjacent applications in our review as they demonstrate technical approaches that could potentially be adapted for VP generation. However, their evaluation metrics will not be discussed in Section 4.

### 4.1 Unconditional Generation

Unconditional generation refers to the synthesis of anatomical structures solely from learned distribution patterns, without explicit control parameters. These models aim to capture the underlying distribution of anatomical shapes and generate diverse, anatomically plausible examples that reflect population-level variations. The primary goal is to produce realistic samples that maintain statistical consistency with the training population while exhibiting natural variability.

#### 4.1.1 Unconditional VAE-based Models

Danu et al. [2019] tested the generation ability of both conventional VAE and GAN in creating synthetic blood vessel surfaces. They developed a voxelization technique that converts the surface meshes into 3D voxels that can be processed by convolutional neural networks. Two key limitations acknowledged are the use of simplified training data and basic architectures; they only generate short segments of vessels with simple topology, thus the models might perform differently with more complex real-world vessel structures like cerebral aneurysms or vessel bifurcations, and the conversion from meshes to voxels might also lead to the loss of topology. The evaluation is also constrained to qualitative visual assessment. Since the methods use conventional VAE and GAN, we do not introduce the architecture again in this section.

Beetz et al. [2022b] introduced a multi-domain VAE framework that jointly models 3D biventricular anatomy from MRI as point clouds and electrophysiology from ECG data within a shared latent space. The model architecture consists of three main components that share a common latent space: 1. Two point cloud branches that handle both healthy and pathological cardiac anatomy at End-Diastolic and End-Systolic phases. They used an extended Point-Net++ [Qi et al., 2017b] architecture for encoding and a Point Completion Network [Yuan et al., 2018] for decoding 3D anatomical structures. The decoder can output both coarse and dense representations of the anatomy; 2. A time-series branch that processes ECG data using convolutional and fully connected layers for both encoding and decoding. A shared latent space that enables joint representation of 3D anatomy and ECG signals is an innovative aspect of the model; however, the ECG signal does not provide any additional control to the anatomy generation. Additionally, the specific types of cardiac pathologies included in the training data are not specified, which limits interpretability and reproducibility.

Kalaie et al. [2024] proposed an unsupervised geometric deep learning framework called ASMG (Attention-based Shape Matching and Generation) for generating virtual left ventricle structures from different-sized training shapes. The framework consists of two main components: an attention-based shape-matching mechanism and a generative model based on *β*-VAE. The shape-matching component employs a variational graph auto-encoder network that captures both local and global structural information in the shapes by computing nodal embedding vectors in the latent space, which are then used to determine correspondences between vertices of different shapes through an attention mechanism. For the generation component, the authors implement a *β*-VAE framework that learns a probabilistic distribution from the structurally normalised shapes. The model uses soft attention to focus on relevant shape features and transforms them into a template domain, allowing for consistent generation across different-sized input shapes.

VesselVAE is a model that generates 3D blood vessel geometries using a recursive variational neural network architecture proposed by Feldman et al. [2023]. It follows the basic structure of the VAE network to handle the hierarchical tree representation of blood vessels. The model does not process the 3D vessel meshes directly; instead, the meshes are converted into binary tree representations. Each vessel segment is parameterised by a central axis with ordered points in Euclidean space and an associated radius. The network takes this binary tree structure as input, where each node represents a vessel segment containing coordinates and radius information. The Encoder transforms the tree structure into a hierarchical encoding on a learned manifold through a depth-first traversal, while the decoder determines whether a node should be a leaf or have bifurcations and reconstructs the geometric attributes of each node. Three auxiliary networks are implemented before and after the bottleneck of the network to shape the distribution of the latent space and decode it. Although the method generates tree-like bifurcations, it is limited in capturing complex vascular topologies, and the final meshes—constructed via post-processing of synthetic centerlines—often contain self-intersections. Future improvements could focus on direct mesh generation to better preserve geometric and topological realism.

Gaggion et al. [2025] introduced HybridVNet, a multiview hybrid graph convolutional network that combines standard VAE with spectral graph convolutions for direct volume-to-mesh extraction from cardiovascular MRI. The model combines traditional convolutional neural networks with graph convolutions to handle both surface and volumetric meshes efficiently. The hybrid encoder processes multiple cardiac MRI views - one 3D short-axis view and three 2D long-axis views - using separate convolutional branches. These branches are concatenated to create a joint latent space representation. The decoder follows a variational approach, sampling from a multivariate Gaussian posterior to generate the mesh.

Dou et al. [2025] proposed a generative shape compositional framework to generate virtual cardiac populations. The cardiac anatomy is composed of the Left Ventricle (LV), Right Ventricle (RV), Left Atrium (LA), Right Atrium (RA), and Aorta (Ao). The framework comprises two key components: a part-aware generative model and a spatial composition network. The part-aware generative model was implemented in two distinct approaches. The Independent Generator utilises separate gcVAEs for each cardiac structure, and each structure has its own dedicated latent space representation. The Dependent Generator employs a single multichannel VAE that learns shared latent representations across all cardiac structures simultaneously. To compose these individually generated parts into complete heart assemblies, they developed a spatial composition network that operates in two stages. First, an affine registration module estimates initial transformations to roughly align the generated parts. This is followed by a non-rigid registration component that refines the alignment by estimating local deformations, particularly at the boundaries between adjacent structures. They introduced an innovative concept of VC to broaden the diversity of the generated virtual populations. However, this model requires a fixed mesh topology as the input, and minor topology defects like intersections between adjacent structures might occur in generated samples.

For the non-generation models, Seale et al. [2024] presented an approach to modelling cardiac anatomy using a *β*-VAE architecture that can reconstruct and predict multiple phases of the cardiac cycle as point clouds from a single input phase. Based on previous works [Beetz et al., 2022b,a, 2023, 2024], they introduced a method to handle latent space representations by using a linear transformation that creates separate but comparable latent spaces for the End-Diastolic and End-Systolic phases. The researchers developed two complementary networks: Network 1, which uses a complete ED point cloud as input, and Network 2, which uses a complete End-Systolic point cloud as input. Both networks are trained to reconstruct and predict point cloud representations for both End-Diastolic and End-Systolic phases. However, the authors only performed qualitative visual evaluations regarding the generation ability of the model.

Cerrolaza et al. [2018] proposed REC-cVAE as a 3D fetal skull reconstruction architecture. The base REC-cVAE employs a multi-branch CNN architecture combining 3D convolutional filters for skull processing with view-specific 2D filter banks for standard ultrasound planes (coronal, sagittal, and axial). Building upon this, they proposed HiREC-cVAE, a hierarchical framework with a three-level nested latent space and cascaded conditional blocks, to progressively refine the geometric features. This hierarchical approach to feature refinement provides a potential mechanism for multi-scale control over anatomical variations in virtual population synthesis. The multi-branch architecture demonstrates one method to incorporate different anatomical viewpoints, which may contribute to maintaining anatomical consistency in generated populations.

Biffi et al. [2018] present a 3D VAE designed to classify cardiac disease states by learning interpretable anatomical features directly from the segmented LV myocardium in cardiac MR images. The architecture consists of an encoder network, a decoder network, and a prediction network. The encoder network compresses these 3D segmentations into a 64-dimensional latent space, while the decoder network then reconstructs the original segmentation. Simultaneously, the prediction network, implemented as an MLP, takes the mean vectors from the latent space and classifies the cases as either healthy or Hypertrophic Cardiomyopathy. Their method of learning interpretable anatomical features in the latent space suggests a possible approach for controlling anatomical variations while maintaining medical relevance in generated samples. They further proposed a VAE-based approach for cardiovascular geometry that combines 2D and 3D processing streams in a unified architecture [Biffi et al., 2019]. The model employs dual encoders: a 2D CNN processing multi-view cardiac images (two longaxis and one short-axis views) and a 3D CNN encoding ground truth segmentations during training. A feature fusion network integrates information from both streams before encoding it into a latent space characterised by mean and variance vectors. The architecture is optimised using a composite loss combining the Dice score for shape accuracy and KL divergence for latent space regularisation. This dual-stream architecture presents a framework for combining different types of geometric information. The feature fusion approach could serve as a basis for integrating multiple anatomical constraints during the generation process.

Abdi et al. [2019] proposed two approaches (deterministic and probabilistic) for jaw reconstruction surgery using VAE. The model architectures centre around a V-Net with four down/up transitions. The networks take a 3D binary map with a missing mandible segment as input and output a 3D probability map. The deterministic one maps the final four features using 2-kernel convolutions and sigmoid activation, while the probabilistic one concatenates the feature maps and processes them with additional convolutional layers. To handle multiple valid reconstruction possibilities, a target-weighted variational objective is introduced, and a voxel-weighted Dice loss is added to prioritise the dissected region while maintaining contextual consistency. While the models can generate multiple valid shapes, they are trained on a small dataset, and the presence of teeth in the generated samples is inconsistent. The authors evaluated this non-deterministic model using a deterministic approach by comparing the generated virtual jaws with the ground truth, which raises questions about whether the model’s full generative capacity was adequately assessed. Their target-weighted variational objective provides a practical mechanism for modelling anatomical variability in population synthesis. The voxel-weighted loss formulation enables region-specific anatomical constraints during generation.

#### 4.1.2 Unconditional GAN-based Models

Wiesner et al. [2019] proposed to create synthetic 3D leukaemia cell shapes using 3D GAN. The model is based on the conventional GAN and extended to handle one additional data dimension. They replace the sigmoid function with a hyperbolic tangent in the last layer of the generator for a more stable performance. This method focuses on the shapes of single leukaemia cells. Therefore, its performance on data with more complicated topologies cannot be guaranteed.

Romero et al. [2021] investigated three approaches for generating synthetic aorta anatomies, along with machine learning surrogates to improve efficiency. The first approach uses non-parametric bootstrapping sampling, which generates new feature vectors by randomly selecting components from observed values in the original small dataset. The second method employs parametric sampling using either multivariate Gaussian or uniform distributions. While Gaussian sampling maintains statistical properties similar to bootstrapping, uniform sampling provides broader coverage of the feature space but with lower acceptance rates. The third approach utilises a GAN trained on the reference cohort. The researchers used the phenotype classification defined by the previous study [Schaefer et al., 2008, Craiem et al., 2011, Casciaro et al., 2014, Bruse et al., 2016, Liang et al., 2017, Sophocleous et al., 2018], which categorizes aortas into three distinct classes based on the relationships between three key measurements, namely the radius of the Sinuses of Valsalva (SoV), the Pulmonary Artery (PA), and the Mid-Ascending Aorta (MA). The data-driven cohort generation was trained on the original 26 patient samples, and the clinically-driven generation was trained on the larger synthetic cohorts corresponding to each phenotype. Although the researchers only used the original GAN and trained on a small dataset, they performed comprehensive evaluations, demonstrated the trade-off between different generation approaches, and validated their ability to produce clinically meaningful synthetic cohorts efficiently.

For the non-generation methods, Sulakhe et al. [2022] proposed CranGAN, a point cloud-based conditional GAN architecture for defective skull completion. The generator adopts an autoencoder structure with a PointNet-based encoder [Qi et al., 2017a]. The CT scans of the defective skulls are converted to 3D meshes and sampled uniformly to point clouds, then they will be sent to the architect for training. Gu and Gao [2023] presented a multi-stage architecture integrating GANs with LSTM and ResNet networks for tumour shape modelling. The framework processes 2D CT slices through a sequential pipeline: initial lung segmentation using snake optimisation, tumour isolation via Gustafson-Kessel (GK) clustering, and finally, 3D reconstruction. The core generative component combines VGGNet for 2D feature extraction, LSTM for temporal sequence compression, and a GAN for 3D shape synthesis. Two works similar to Gu and Gao [2023] that also incorporate LSTM with GAN are Rezaei and Ahmadi [2023] and Hong et al. [2023]; they also use GK clustering as the lung tumour segmentation method. The main difference between these two works is the usage of VGG instead of ResNet. The integration of temporal sequence processing with spatial feature extraction presents a potential approach for maintaining consistency across different anatomical cross-sections in population generation.

The Hierarchical Shape-Perception Network (HSPN) proposed by Hu et al. [2024a] represents a novel approach combining encoder-decoder architecture with GAN-based adversarial learning for point cloud generation. The architecture integrates multiple PointNet++ encoding blocks with Attention Gate Blocks (AGBs) for hierarchical feature extraction and aggregation. A key innovation is the incorporation of structural consistency decoding blocks that ensure anatomical plausibility in the generated shapes. The structural consistency mechanisms could be adapted for maintaining anatomical relationships in generated populations.

SG-GAN [Hu et al., 2025] is a two-stage GAN framework combining stereoscopic-aware graph convolutions with a fine-grained up-sampling mechanism to generate high-density brain point clouds from a single 2D MRI image. The model starts with a novel free-transforming module (FTM), which employs parameter-free self-attention mechanisms to efficiently extract features from input MRI images without adding computational overhead. This can be beneficial for real-time surgical applications. The Stage-I GAN employs a tree-structured graph convolutional network to outline the basic anatomical structure, and the Stage-II GAN then performs intelligent upsampling to generate the final high-density point cloud.

Hu et al. [2021] proposed to reconstruct 3D brain shapes from single 2D MRI images using a tree-structured graph convolutional network. The key innovation lies in its architectural design, which combines graph convolutions with a tree-structured generative process. The model has a ResNet-based encoder, a tree-structured GCN generator, and a discriminator. Each GCN block in the generator incorporates two key elements: a loop term for feature propagation and an ancestor term for maintaining structural relationships. This design allows the model to progressively expand a single point into a complete point cloud through controlled branching operations.

The Dental Mesh Completion (DMC) model [Hosseinimanesh et al., 2023] presents an innovative end-to-end approach for generating dental crown meshes directly from 3D scans. The architecture consists of three main components working in concert to produce crown reconstructions. The first component is a transformer-based encoder-decoder that uses a dynamic graph convolution network (DGCNN) to group input points into feature vectors representing local regions. These features are then processed through the encoder’s geometry-aware blocks and self-attention layers to capture both local geometric relationships and long-range dependencies. The decoder uses self-attention and cross-attention mechanisms to reason about the crown structure based on the encoded features. The second key component is a folding-based decoder that transforms a canonical 2D grid into the 3D surface of the crown points. The final component is a differentiable Poisson surface reconstruction (DPSR) layer that converts the point cloud into a complete mesh.

Chafi et al. [2024] proposed a new method for automatically generating dental crown bottoms using GANs, bypassing the need for manual manipulation by dental technicians. The deep learning model adapts the SP-GAN architecture for crown bottom generation. The approach uses a sphere of 2048 points as a global before enable unbiased spatial deformation, combined with a Gaussian function that provides random latent code sampling. The network architecture consists of two main components: a generator that creates dense correspondences between the global prior and generated point clouds using part-wise interpolation, and a discriminator that evaluates the authenticity of generated shapes. The model incorporates graph attention modules connected to adaptive instance normalisation for enhanced detail preservation.

#### 4.1.3 Unconditional Flow-based Models

Peng et al. [2024] proposed an approach to generating virtual populations of 3D cardiac anatomies (LV endocardium, LV epicardium, RV endocardium) using the Snowflake-Net architecture [Xiang et al., 2022]. The model employs a PointNet encoder to progressively capture features from input point clouds representing cardiac structures, ultimately producing a compressed latent representation. The decoder network then generates high-quality anatomies through a coarse-to-fine strategy using Snowflake Point Deconvolution modules. To enable the effective generation of new cardiac anatomies, the researchers incorporate a normalising flow component that transforms a simple Gaussian distribution into a more complex one capable of capturing the intricacies of cardiac shape variation.

#### 4.1.4 Unconditional Diffusion-based Models

TrIND [Sinha and Hamarneh, 2024] introduced a novel two-stage approach for representing and generating anatomical tree structures. The first stage involves learning per-sample Implicit Neural Representations (INRs), while the second stage employs a diffusion model to capture the distribution of these representations. For the first stage, the authors represent each anatomical tree as a neural occupancy field using an MLP. Unlike previous approaches that share parameters across datasets, TrIND optimises an INR separately for each tree sample. The network architecture consists of a fully connected network with ReLU activations, trained to minimise the L2 distance between predicted and ground truth occupancy values. In the second stage, TrIND trains a transformer-based diffusion model on the space of INR parameters. For generations, the model employs DDIM sampling to produce novel INR parameters by gradually denoising random noise. They performed the evaluations on multiple datasets but they failed to specify the training-testing split.

Kadry et al. [2024a] uses a two-stage approach where cardiac label maps are first encoded into latent space using a VAE, then edited using a DDIM through two methods: (1) perturbational editing, which applies partial noise and denoising for scale-specific variations, and (2) localised editing, which uses masked sampling for region-specific changes while preserving other areas. The process involves encoding a seed image, applying controlled noise or masked sampling, and then decoding back to anatomical space. They further proposed another diffusion model that implements a multi-stage generation workflow that begins with encoding coronary MRI images into a latent space using a topologically regularised VAE [Kadry et al., 2024b]. The key innovation lies in its specialised conditioning and guidance mechanism for anatomical generation. Unlike standard LDMs that typically use single-type conditions, this model incorporates three parallel conditions: morphological features capturing local tissue characteristics, skeletal features controlling global artery branching patterns, and stochastic latent codes from DDIM inversion.

For the reconstruction tasks, the DMCVR [He et al., 2023] takes 2D cardiac cine MRI slices from short-axis views as input and processes them through three main neural network components to reconstruct 3D LV and RV structures. Initially, the slices pass through a global semantic encoder that captures high-level image features and simultaneously through a regional morphology network that specifically focuses on cardiac structures (LVC, LVM, and RVC). These two networks generate semantic and morphological latent codes, respectively. The model then uses a DDIM for image reconstruction. Finally, these generated slices are combined with the original ones to create a high-resolution 3D cardiac volume.

Jayakumar et al. [2023] applied an atlas-building network process to 3D myocardium training images to learn and generate a mean shape (atlas). Then, they used a diffusion model guided by three key elements: the shape features from the atlas, the input sparse 2D slices, and a stochastic latent code to deform the atlas to a specific variant. Although this is designed for reconstruction tasks, the deformation field can be altered to satisfy the virtual population generation demands.

#### 4.1.5 Unconditional DeepSSM-based Models

Inspired by DeepSDF [Park et al., 2019], Gasparovici and Serban [2024] proposed a deep learning approach for generating synthetic aortic valve meshes for ISTs. The model employs an auto-decoder neural network architecture that represents 3D shapes as zero-level sets of a neural signed distance field. The network takes as input both 3D coordinates and learned shape embedding vectors, which encode the geometric features of each shape. These embedding vectors serve a similar role to the shape parameters in traditional SSMs but can capture more complex, nonlinear variations. However, a Marching Cubes meshing algorithm [Lorensen and Cline, 1987] is required to convert the SDF to the final mesh, and the evaluations are not comprehensive enough in terms of fidelity.

### 4.2 Conditional Generation

#### 4.2.1 Conditional VAE-based Models

Huang et al. [2024] presented a unified framework for modelling and generating rib cross-sectional shapes while accounting for population variations and individual characteristics. The framework consists of three integrated components. At its core is a cVAE that learns to represent rib shapes while disentangling two key types of variation: inter-class differences (based on rib number and sex) and intra-class variations (individual differences within each category). Finally, random forest regressors correlate the learned shape representations with anthropometric measurements like age, height, and weight.

Both CHeart [Qiao et al., 2024] and MeshHeart [Qiao et al., 2025] utilised VAE structures to generate 3D+t cardiac mesh sequences conditioned on clinical factors. However, CHeart builds upon a *β*-VAE framework with a specialised temporal module using LSTM networks, focusing on learning a structured latent space that separates motion features from structural features. They further proposed MeshHeart to model and generate 3D+t cardiac mesh sequences while accounting for clinical factors like age, sex, weight, and height. The model’s architecture integrates several key components working in concert: a mesh encoder processing the input cardiac mesh sequence, a condition encoder handling clinical parameters simultaneously, a Transformer encoder incorporating distribution parameter tokens to capture the statistical properties of the cardiac motion patterns, and a Transformer decoder and mesh decoder producing the final 3D+t cardiac mesh sequence. The model employs a reparameterization technique to generate latent vectors before the decoder.

For other tasks, Foti et al. [2020] presented a method for intraoperative liver surface completion from the limited view available during surgery for surgical guidance using a graph VAE. During training, the model is trained on 50 complete and standardised liver meshes from CT scans, and the model learns on this standardised mesh representation using graph convolutions that operate directly on the mesh structure. For inference, they provide a non-rigid shape completion method that enhances real-time intraoperative visualisation and guidance by finding the best match for the point cloud of the partially visible liver during the surgery. However, the method is only trained and tested on a small dataset of 50 samples; the stability of this model might require further validation.

Biffi et al. [2020] proposed a hierarchical VAE framework based on Ladder VAE, in which the higher levels capture global anatomical features of the LV and hippocampus meshes while lower levels encode local details. This architecture combines generative capabilities with clinical classification (hypertrophic cardiomyopathy and healthy for LV, AD and healthy for hippocampus) through its highest latent level as additional controls.

Beetz et al. [2022d] proposed to use a variational mesh autoencoder to process the cardiac anatomy of different phases (End-Diastolic/End-Systolic) through direct mesh operations, utilising graph convolutions and hierarchical mesh sampling. This method combines graph convolutional layers using Chebyshev polynomial approximation with mesh pooling based on quadric error minimisation, encoding shapes into a compact 16-dimensional latent space. They then introduced a conditional *β*-VAE framework utilising PointNet++ and Point Completion Network (PCCN) architectures to generate subpopulation-specific biventricular anatomy models directly from point cloud data [Beetz et al., 2023]. The encoder takes sparse, misaligned point clouds as input, where each point is represented by a 4-dimensional vector containing spatial coordinates and a class label identifying the cardiac substructure (LV endocardium, LV epicardium, or RV endocardium). The encoder processes this through multiple PointNet-style convolutional blocks and pooling operations to create a latent space representation. The decoder has a two-stage design. The first stage generates a coarse point cloud, and the second stage uses a FoldingNet-inspired approach that initialises patches corresponding to points in the coarse cloud.

#### 4.2.2 Conditional GAN-based Models

Wolterink et al. [2018] presented a framework based on WGAN for generating anatomically accurate blood vessel geometries through a data-driven methodology. In their approach, the system operates on a latent probability distribution from which random vectors are sampled to initiate the generation process. The model’s representation of vessels is the 1D parametric central line, where each point is characterised by a 4-dimensional vector encoding 3D spatial coordinates and vessel radius along the centre line. The framework incorporates conditional generation capabilities through an attribute vector system, building upon the cGAN framework. This is achieved by augmenting both the generator and discriminator networks with additional input channels corresponding to the desired attributes (length, left or right coronary tree). However, no bifurcations are modelled in this method, and the transformation from the central line to the coronary vessel shapes by post-processing is required.

Hwang et al. [2018] proposed an approach to automating dental crown design using a GAN. The core architecture builds upon the pix2pix conditional GAN framework, using a U-Net generator with skip connections and a convolutional discriminator. The model is enhanced with conditioning on both the prepared jaw and opposing jaw scans to handle the specific requirements of the dental crown design. It introduces novel functionality constraints through a histogram loss that captures the natural spatial statistics between opposing teeth, ensuring proper biting and chewing functionality. However, this work is limited to a single tooth and requires conversion between 2D and 3D representations.

GANs are also applied to prediction tasks for tumours and hearts. To predict tumour growth, a stacked conditional GAN architecture (GP-GAN) is proposed by Elazab et al. [2020]. The model consists of *n* − 1 sequential GANs, where *n* represents the number of time points. Each generator processes a T1 MRI volume alongside semantic labels (tumour boundary, white/grey matter, cerebrospinal fluid), while its corresponding discriminator validates the generated tumour boundary against the next time point’s ground truth. Yuan et al. [2020] introduced a CGAN framework specifically tailored to reconstruct dental occlusal surfaces, conditioned on the spatial relationship with the opposing tooth. Each sample of the training set consists of depth maps of the tooth, the opposing tooth, and the technician-designed crown. The occlusal groove filter network is trained and frozen first, and the generator uses this frozen filter along with L1 loss to generate crowns. Finally, perceptual loss is introduced to fine-tune the network. In DCPR-GAN [Tian et al., 2022], they proposed a two-stage GAN architecture to progressively refine the dental crown reconstruction. In the first stage, a conditional GAN learns the inherent relationship between the defective tooth and target crown, focusing on restoring proper occlusal relationships. This stage utilises a DGCNN encoder to capture local geometric features and an attention module to generate correspondence mappings. The second stage introduces an improved conditional GAN that incorporates two key components: an occlusal groove parsing network (GroNet) and an occlusal fingerprint constraint. These additions help ensure the generator produces anatomically accurate surface details. The GroNet is pre-trained on Stage-I results and remains fixed during Stage-II training to provide consistent groove feature extraction.

For cardiac anatomy, Qiao et al. [2022] proposed a conditional GAN model for ageing heart synthesis that takes a cardiac anatomy image and target age as input, and generates the predicted anatomy at that age. The model uses an encoder-decoder architecture with a condition mapping network to integrate age and gender information, trained using cycle-consistent and adversarial losses.

#### 4.2.3 Conditional Flow-based Models

Dou et al. [2023] proposed a conditional flow variational autoencoder (cVAE-NF) that combines two key elements to enable the controlled synthesis of anatomical structures. First, it incorporates conditioning information through demographic and clinical measurements (like gender, age, weight, blood cholesterol, etc.) that guide the generation process. The triangular mesh representation of cardiac structures, along with 14 clinical/demographic covariates, is encoded through a graph convolutional encoder. Second, it employs normalising flows in the latent space to transform the initial Gaussian distribution into a more flexible multi-modal distribution. The conditioning is implemented in two ways: by scaling the hidden representations in the encoder based on the covariates and by concatenating the covariates with the latent variables before decoding.

SDF4CHD [Kong et al., 2024] is a deep-learning approach for modelling cardiac anatomies with congenital heart defects (CHDs). The model consists of three key components working together: a type representation module, a diffeomorphic shape representation module, and a correction module. The type representation module leverages CHD diagnostic information to predict type-specific cardiac geometries using SDFs. It employs multilayer perceptrons to learn implicit representations of seven cardiac structures, including the myocardium, blood pools, and major vessels. The module takes CHD diagnosis vectors as input and outputs signed distance values that capture the unique topological features associated with different CHD types. The shape representation module then handles patient-specific variations while preserving the CHD-specific anatomical features. It uses NODEs to learn invertible flow between the type-specific templates and patient-specific shapes. To capture additional anatomical variations not fully described by the CHD types, the correction module provides per-point adjustments to the deformed geometries.

#### 4.2.4 Conditional Diffusion-based Models

Kuipers et al. [2024] also utilised the diffusion model in vessel generation. They introduced a conditional set-diffusion approach that generates vessel trees by learning point cloud representations conditioned on the presence of M1 occlusion, using cross-attention mechanisms for incorporating the conditional information. The vessels are parameterised as point coordinates with their features (radius, vessel segment labels). Initially, the model samples a noisy point cloud and processes it alongside condition labels specifying attributes. Next, the reverse diffusion process iteratively denoises this point cloud to form an unordered set of vessel centreline points. Finally, a post-processing sequencing algorithm orders these points into coherent vessel segments (ICA, ACA, M1, and M2), handling bifurcations and merging segments appropriately. However, this method occasionally fails on M2 segments, and the generation requires a post-processing step. Although centrelines are important for understanding vessel topology and path planning, the authors should plot a full 3D vessel geometry instead of only a centreline visualisation. It could be a more solid work if the samples generated are shaped and validated via simulations, as there are related simulation approaches like those presented by Miller et al. [2021].

Zhang et al. [2024a] proposed a conditional diffusion model architecture for dental crown point cloud generation that operates through several interconnected components and stages. The model begins with an encoder that processes input point cloud data into the global representations. This encoder produces both mean and standard deviation parameters that define the shape of the latent distribution. The model passes the latent features through an MLP that maps them to multiple part-level vectors, each representing different regions of the dental crown. These part-level representations are then processed through a cross-attention block, where the global shape latent serves as queries while the part-level vectors act as keys and values. The diffusion process itself follows a forward-reverse framework. Each step of the reverse process is guided by the part-level and shape latent representations to denoise the input point cloud.

#### 4.2.5 Conditional DeepSSM-based Models

Xia et al. [2022] presented MCSI-Net (Multi-Cue Shape Inference Network), an innovative deep learning architecture designed for generating accurate 3D cardiac models from CMR images. The model incorporates both imaging and demographic patient data to create comprehensive four-chamber cardiac generations. The core architecture consists of two primary components: MMF-Net for shape parameter prediction and Loc-Net for spatial transformation estimation. MMF-Net processes input from short-axis (SAX) CMR views, long-axis (LAX) CMR views from different perspectives, and patient metadata, including demographics and clinical measurements. The imaging feature is extracted through a ResNet-based encoder, and the patient data is processed through an MLP. The extracted features are then concatenated and used to predict shape parameters in a learned PCA space.

#### 4.2.6 Other Models

Sørensen et al. [2024] introduced a conditional generative model that uses neural distance fields to represent and generate cardiac anatomical sequences while incorporating clinical demographic information. Instead of using the common generative models mentioned before, the model employs an Auto-Decoder architecture that learns two separate latent spaces - one capturing clinical demographic factors and the other representing individual variations. A spatio-temporal neural distance field maps space-time co-ordinates to SDFs, which creates decision boundaries that define the cardiac surface geometry across time. The model takes clinical demographic data (gender, age, and systolic blood pressure) as input and encodes it into a clinical demographic latent vector using a neural network. Additionally, it learns a residual latent vector for each training sequence to capture individual variations not explained by demographics. However, the required test time optimisation could be more time-consuming according to the authors.

## 5 Evaluation Metrics for Synthetic Anatomical Data

Evaluating the quality of synthetic anatomical data (in this case, VP) is crucial to ensure that it is both anatomically realistic and functionally useful. Without robust evaluation metrics, it becomes challenging to assess how well VP data preserves the anatomical fidelity, variability, and clinical relevance found in real-world datasets. This section presents key quantitative and qualitative metrics for evaluating synthetic anatomical data across multiple dimensions—such as *fidelity, diversity, utility, and generalizability*—offering a comprehensive framework for validating generative models in medical applications. [Hernadez et al., 2023, Alaa et al., 2022].

### Fidelity

The fidelity dimension evaluates how well synthetic data preserves the statistical distributions of single variables while also maintaining the complex relationships between multiple variables [Murtaza et al., 2023].

### Utility

The utility assessment focuses on whether synthetic data can effectively stand in for real data in downstream tasks [Hernandez et al., 2022]. This involves comparing how well machine learning models perform when trained on synthetic versus real data.

### Generalizability

Generalizability quantifies the extent to which a generative model overfits (copies) training data [Alaa et al., 2022].

### Diversity

Diversity represents how comprehensively synthetic data encompasses the patterns and variations found in real data [Alaa et al., 2022].

These dimensions often involve inherent trade-offs [Hernadez et al., 2023, Alaa et al., 2022]. Maximising utility may reduce privacy protections, while strict privacy requirements could negatively impact resemblance and utility; such trade-offs also exist. On the contrary, maximising fidelity and diversity may enhance the representativeness of the synthetic data, but could also reduce utility or generalisation. Therefore, using multiple complementary metrics within each dimension is recommended to obtain a comprehensive assessment of synthetic data quality. Our first-level evaluation classification of the metrics is based on this taxonomy.

For the second-level classification, we can classify evaluation metrics into two fundamental categories: *shape-based* and *feature-based*. Shape-based metrics measure the similarity between generated and real anatomical samples by directly comparing their geometric representations. These metrics operate on the raw anatomical representations themselves and calculate various distance or similarity measurements between them. Feature-based metrics evaluate generated anatomies by comparing derived properties, biomarkers, or clinical measurements rather than the raw geometric representations. These metrics assess whether the generated anatomies maintain clinically relevant characteristics and relationships found in real populations.

### 5.1 Fidelity

For fidelity, the shape-based metrics include 1st nearest neighbour accuracy (1-NNA), coverage score (COV), divergence metrics, minimum matching distance (MMD), specificity, and statistical tests. The feature-based metrics include anthropometric analysis, clinical acceptance rate, and interpolation studies.

#### 5.1.1 1st Nearest Neighbour Accuracy

The 1-NNA is a classification-based evaluation metric evolved from k-nearest neighbour algorithms [Zhang, 2016]. It assesses whether two samples are drawn from the same cohort by measuring how well a simple nearest neighbour classifier can distinguish between them.

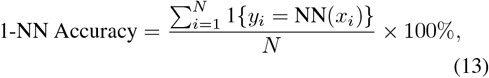

where *N* is the total number of samples, *y*_*i*_ is the true label of the i-th sample, and *NN* (*x*_*i*_) is the predicted label of the nearest neighbour for *x*_*i*_.

This metric provides insights into the quality and distinctiveness of generated samples by evaluating whether they maintain characteristics similar to the real data distribution while remaining appropriately distinguishable. The ideal score is 50%, indicating that real and generated samples are indistinguishable from each other. This metric has been employed in recent research evaluating generative models, as demonstrated by [Sinha and Hamarneh, 2024, Peng et al., 2024].

#### 5.1.2 Divergence

Divergence metrics are used to quantify the dissimilarity or distance between two probability distributions. In the context of generative models for medical anatomical structures, these metrics are essential for evaluating how well the distribution of generated samples matches the distribution of real anatomical data. This is crucial for applications like ISTs or VP generations, where we need to ensure that generated anatomies represent realistic variations seen in actual patient populations. Three commonly used divergence metrics in medical imaging applications are the Kullback-Leibler Divergence (KLD) [Kullback and Leibler, 1951], Earth Mover’s Distance (EMD) [Rubner et al., 2000], and Jensen-Shannon Divergence (JSD) [Lin, 1991].

KLD quantifies how one probability distribution differs from another. For continuous probability distributions P and Q, it is defined as:

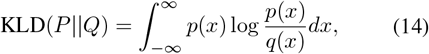

where *p*(*x*) is the probability density function of the real data distribution, *q*(*x*) is the probability density function of the VP distribution, and *x* represents a single data point or sample in the space where both real and VP distributions exist.

EMD is a special case of the Wasserstein Distance (WD), specifically the Wasserstein-1 distance applied to discrete probability distributions. As formulated in Equation 15, *P* represents the probability distribution of real data (e.g., training images or text), *Q* is the distribution of samples produced by your generative model, *x* denotes a point from the real distribution, *y* denotes a point from the generated distribution, *d*(*x, y*) is the “ground distance” measuring dissimilarity between individual points *x* and *y* (like pixel-wise distance for images), *γ*(*x, y*) represents the transportation plan determining how probability mass moves from *x* to *y*, Π(*P, Q*) is the set of all possible joint distributions with marginals *P* and *Q*, and inf indicates the minimum transportation cost across all possible plans.

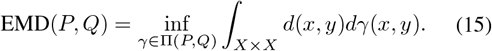

JSD measures the similarity between two probability distributions while being symmetric and bounded. For distributions P and Q, it is defined as:

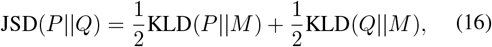

where 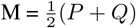.

The above divergence metrics are also used to compare the distributions of the *derived biomarkers*, which will be further introduced in the feature-based anthropometric analysis later.

#### 5.1.3 Maximum Mean Discrepancy

The Maximum Mean Discrepancy [Gretton et al., 2007] is a quantitative metric that measures how well a generative model can capture the statistical properties of a real data distribution. For geometric generation tasks, MMD evaluates the distance between the distributions of generated and real samples to assess both the quality and diversity of the synthesised outputs. A lower MMD score indicates that the generated distribution better matches the real data distribution, suggesting higher quality geometric synthesis. The metric provides a principled way to quantify how well generative models can capture the underlying distribution of medical geometries. It can be formulated as:

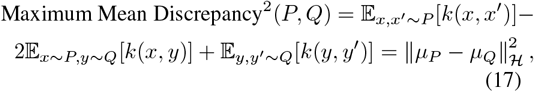

where *P, Q* are two probability distributions being compared; *k*(·, ·) is the kernel function (typically a Gaussian RBF kernel: 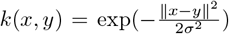 𝔼 is the expectation operator; *x, x*^*′*^ are independent random variables drawn from distribution *P* ; *y, y*^*′*^ are independent random variables drawn from distribution *Q*; *µ*_*P*_, *µ*_*Q*_ are mean embeddings of the distributions *P* and *Q* in the reproducing kernel Hilbert space (RKHS) ℋ ∥ϻ∥; _ℋ_ is the norm in the reproducing kernel Hilbert space.

In real-world applications, we rarely have access to the complete probability distributions *P* and *Q*. Instead, we typically have finite samples drawn from these distributions (e.g., real data versus generated data). The empirical MMD allows us to compute the discrepancy using only these available samples, which can be formulated as:

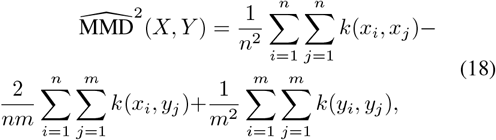

where *X* = {*x*_1_, *x*_2_, …, *x*_*n*_} is a sample set of size *n* drawn from distribution *P* ; *Y* = {*y*_1_, *y*_2_, …, *y*_*m*_} is a sample set of size *m* drawn from distribution 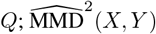 is the empirical estimation of the squared Maximum Mean Discrepancy (MMD) using samples *X* and *Y* ; *n* and *m* are the sample sizes of sets *X* and *Y* respectively.

This metric has been adopted in anatomical geometry generation applications. Beetz et al. [2022b] used Maximum Mean Discrepancy to evaluate both standalone ECG generation and joint ECG-anatomy generation quality.

#### 5.1.4 Minimum Matching Distance

Minimum Matching Distance (MMD) is a metric used to quantify the similarity between two point sets from the ground truth set and the VP set by finding the optimal correspondence or matching between their points. It can be calculated using CD or EMD. Thus the formulation of MMD can be expressed as:

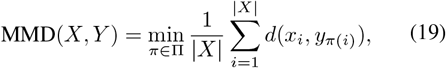

in which Π is the set of all possible permutations of indices, *π*(*i*) represents a specific matching from points in *X* to points in *Y*, and *d*(·, ·) is a distance metric between individual points (CD or EMD).

In the context of dental geometries, Zhang et al. [2024a] employed MMD to assess the distributional alignment between generated and real dental crown point clouds. The metric has also been utilised by Sinha and Hamarneh [2024] and Peng et al. [2024] to evaluate the fidelity of their generated anatomical structures.

#### 5.1.5 Specificity

Specificity measures whether a generative model can generate new shapes that are anatomically plausible and similar to the training examples while avoiding the creation of unrealistic or invalid shape variations. A highly specific model ensures that any newly generated shape instance closely resembles the actual shapes found in the training set.

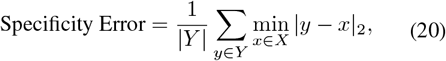

where *Y* is the set of all VP, *X* represents the set of all samples in the real population, |*Y* | is the number of samples in the VP, and |*y* − *x*|_2_ is the distance between a virtual patient *y* and a real patient *x*.

This property is particularly important in medical applications, where generated shapes must maintain anatomical validity [Davies et al., 2010, Styner et al., 2003, Munsell et al., 2008]. The calculation of specificity typically involves computing distance metrics between each generated sample and its nearest neighbour in the training dataset. As evidenced in the literature, researchers have applied various approaches to calculate specificity. For instance, Kalaie et al. [2024] employed multiple distance metrics (HD, ED, and symmetric ED) to evaluate generated Left Ventricle (LV) populations against training samples. Similarly, Kuipers et al. [2024] utilised CD for their specificity calculations, while Dou et al. [2023, 2025] focused on ED measurements between generated cardiac shapes and their nearest neighbours in existing populations.

#### 5.1.6 Statistical Tests

Statistical Tests serve as rigorous tools for evaluating whether generated medical geometries faithfully preserve the distributional properties of real anatomical data. These tests assess whether differences between real and synthetic data distributions are statistically significant, providing quantitative validation of generative models’ ability to capture population-level characteristics. The Kolmogorov-Smirnov Test (KS-Test) [Massey, 1951] is a non-parametric statistical test that measures if two samples are drawn from the same continuous distribution. For two cumulative distribution functions *F*_1_ and *F*_2_, the KS statistic is defined as:

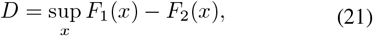

where sup represents the supremum of the set of distances. The larger the value of *D*, the more likely the two distributions are different.

The Mann-Whitney-Wilcoxon Test (MWW) [Wilcoxon, 1945], also known as the Wilcoxon rank-sum test, evaluates whether two independent samples come from populations with the same distribution. For samples *X* and *Y* of sizes *m* and *n*, the test statistic *W* is calculated from the ranks of the combined samples:

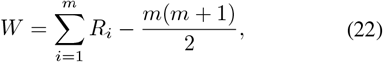

where *R*_*i*_ represents the rank of sample *i, m* is the minimum possible sum of ranks for the first sample group. If *W* is significantly different from zero (either positive or negative), it suggests that one population tends to have higher mass, with some studies further stratifying these comparisons across demographic subgroups. These studies also employed KLD to evaluate how well the learned latent distributions aligned with the target Gaussian distribution values than the other, indicating that the two samples likely come from different distributions.

These statistical tests provide complementary approaches to validating generative models - while KS-tests focus on overall distributional similarity, MWW tests can capture more subtle differences in population characteristics. Together, they enable rigorous validation that generated geometries maintain statistical properties of real anatomical structures. These statistical tests can also be applied to *feature-based* evaluations of the derived biomarkers, which will be further introduced later.

#### 5.1.7 Anthropometric Analysis

Anthropometric and clinical measurement analysis evaluates the generated 3D anatomical shapes by assessing both their geometric properties and derived clinical measurements against real anatomical data. This method determines whether synthetically generated anatomies maintain the correct proportions, dimensions, spatial relationships, and clinically relevant metrics that are essential for medical applications.

The evaluation involves calculating specific geometric parameters and clinical measurements from both real and generated anatomical structures and then comparing their statistical distributions through visualisations like histograms and kernel density plots. The specific measurements analysed vary based on the anatomical structure and its clinical relevance.

This approach has been implemented across various anatomical domains: For vascular structures, Wolterink et al. [2018] analysed blood vessel length distribution, volume measurements, and tortuosity metrics as defined by Bullitt et al. [2003]. Feldman et al. [2023] compared distributions of total length, average radius, and tortuosity between real and synthetic vessel samples, while Kuipers et al. [2024] extended this to include bifurcation angles and employed KS-tests to validate the distributional similarity of vessel characteristics.

In dental applications, Hwang et al. [2018] focused on the distribution of contact points between opposing teeth to evaluate proper occlusion relationships and potential interference. Zhang et al. [2024a] used JSD to effectively quantify the distributional similarity between generated and real dental crown point clouds.

For cardiac structures, Dou et al. [2023] examined left ventricular blood pool volume distributions, while Qiao et al. [2025, 2024] performed more extensive analyses of ventricular volumes, ejection fractions, and myocardial mass, with some studies further stratifying these comparisons across demographic subgroups. These studies also employed KLD to evaluate how well the learned latent distributions aligned with the target Gaussian distribution and used EMD to compare the distributions of various cardiac function metrics between real and virtual populations. Sørensen et al. [2024] analysed left atrial fractional change distributions across different demographic categories. Qiao et al. [2022] also applied EMD for comparing cardiac function metrics.

For cellular structures, Wiesner et al. [2019] conducted a comprehensive geometric analysis examining volume, surface area, roundness, radius measurements, elongation, and rectangularity of synthetic cells compared to real cells, and used KS-tests to compare PCA component distributions between synthetic and real data.

#### 5.1.8 Interpolation Study

Interpolation Study is a technique used to evaluate whether a generative model has learned a continuous and meaningful latent space representation of anatomical structures. This method involves examining how the generated anatomies change when linearly interpolating between two points in the latent space. The effectiveness of interpolation studies has been demonstrated across various anatomical geometry generation applications. For instance, Wolterink et al. [2018] used this approach to verify continuous latent representations of blood vessel geometries. Beetz et al. [2022b] extended this analysis to identify interpretable latent space components controlling specific geometric features of the heart - the first three components were found to govern overall heart size, short-axis tilt, and basic shape, respectively. In the context of Congenital Heart Disease (CHD), Kong et al. [2024] employed interpolation to demonstrate that their model’s latent space could represent the complete geometric spectrum of CHD variations.

This evaluation method serves two crucial purposes in geometric generation: it verifies the continuity of the learned geometric representation and helps identify interpretable features in the latent space, which is essential for the controlled generation of anatomical structures with specific geometric properties.

### 5.2 Utility

Utility refers to the ability of synthetic data to effectively train machine learning (ML) models, producing similar results to those achieved with real data. It is calculated by comparing the Train on Real, Test on Real (TRTR) and Train on Synthetic, Test on Real (TSTR) performance of the same downstream task model [Hernandez et al., 2022]. All metrics for utility evaluation are feature-based.

#### 5.2.1 Clinical Acceptance Rate

The Clinical Acceptance Rate [Romero et al., 2021] is an evaluation metric that assesses the physiological plausibility of generated anatomical structures by measuring the proportion of generated samples that meet predefined clinical and anatomical criteria. This metric combines statistical analysis of anatomical measurements with clinical expertise to ensure the generated structures maintain clinically relevant properties.

The calculation of the clinical acceptance rate involves establishing acceptance criteria through both data-driven and clinically-driven approaches. The data-driven approach typically defines confidence intervals for anatomical measurements using statistical methods. As demonstrated [Romero et al., 2021], these can include multiple tiers of acceptance criteria. We summarised five confidence intervals that have been used in the publications reviewed:

- min */* max: Accepts anatomies with biomarkers falling within the observed minimum and maximum values of the reference dataset [Romero et al., 2021, Kalaie et al., 2024].
- *µ* ±2*σ*: Accepts anatomies with biomarkers within two standard deviations of the mean, covering approximately 95% of a normal distribution. It is based on the *Empirical Rule* [Romero et al., 2021].
- *µ* ± *σ*: Accepts anatomies with biomarkers within one standard deviation of the mean, covering approximately 68% of a normal distribution. It is also based on the *Empirical Rule* but tighter [Kalaie et al., 2024].
- *M* ± 3*B*: Accepts anatomies with biomarkers within three specialised bounds (*B*) from the mode (*M*), containing at least 95% of values in a unimodal distribution according to Chebyshev’s theorem [Romero et al., 2021].
- *M* ± 2*B*: A more restrictive version of *M* ± 3*B* that uses two specialised bounds from the mode instead of three [Dou et al., 2025, Kalaie et al., 2024].

This evaluation method has been implemented in various anatomical contexts. In thoracic aorta analysis, Romero et al. [2021] evaluated 11 anatomical biomarkers using three different acceptance functions and incorporated phenotype classifications based on relationships among the SoV, PA, and MA from Schaefer et al. [2008]. This approach has been adapted by other researchers for different anatomical structures. For instance, Kalaie et al. [2024] applied similar confidence interval designs to evaluate left ventricular volume, while Dou et al. [2025] extended the evaluation to all four cardiac chamber volumes. The final acceptance rate is calculated as the ratio of samples meeting all criteria to the total number of generated samples, providing a quantitative measure of the generator’s ability to produce clinically valid anatomical structures.

#### 5.2.2 Downstream Task

Downstream Tasks evaluation assesses how well VP performs when used for various real-world clinical applications and analyses. This metric evaluates whether synthetic data can effectively substitute for real patient data in supporting important healthcare tasks, particularly focusing on classification and regression tasks.

In classification tasks, the VP is evaluated based on its ability to support accurate clinical categorisation and decision-making. Recent research demonstrates diverse applications of this evaluation approach. For instance, Romero et al. [2021] employed an SVM classifier to predict which phenotype (SoV *>* PA and SoV ≥ MA, SoV *>* PA and SoV *<* MA, or SoV ≤ PA) an aorta belongs to based on its feature vector in PCA space. Beetz et al. [2022b] examined how different combinations of encoded anatomical and ECG data affected classification performance. They used a binary classification approach to identify subjects with at least one pathology related to the cardiovascular system. In Qiao et al. [2025], for each of the six cardiovascular conditions (myocardial infarction, ischemic heart diseases, paroxysmal tachycardia, atrial fibrillation/flutter, hypertension, and cardiac disease), they train separate binary classifiers that predict whether a subject has that specific condition or not.

For regression tasks, the utility assessment examines whether models trained on VP can accurately forecast clinical outcomes or characteristics. Huang et al. [2024] demonstrated this by evaluating their models’ ability to predict various clinical metrics of the ribs, including age, height, and weight, through surface and thickness predictions.

The comprehensive evaluation of downstream tasks provides crucial insights into the practical value of VP generation systems, ensuring that the VP not only maintains statistical similarity to real data but also supports the actual clinical applications it is intended to enable.

#### 5.2.3 Clinical Use Validation

Clinical Use Validation refers to the process of assessing whether synthetic or virtual patient (VP) data can be safely and effectively used within real-world clinical workflows. This involves verifying not only the technical performance of the generative model but also its alignment with regulatory, ethical, and safety standards required in healthcare.

For example, Hwang et al. [2018] noted that their system was being tested for potential production use, which marks a preliminary step toward clinical deployment. However, such statements should be accompanied by robust evidence from clinical trials or evaluations that demonstrate the data’s safety, reliability, and compatibility with medical decisionmaking processes.

To bridge the gap between theoretical model performance and real-world applicability, more comprehensive clinical validation studies are necessary—ideally including stakeholder engagement (e.g., clinicians), prospective testing, and adherence to standards from regulatory bodies such as the FDA.

### 5.3 Generalizability

Generalizability in the context of generative models quantifies the extent to which a model avoids overfitting to training data. In current anatomical generative model literature, this is predominantly assessed through shape-based reconstruction metrics of unseen instances, typically using leave-one-out reconstruction approaches [Davies et al., 2010]. It is important to note that this assessment has been primarily limited to models with deterministic properties in this review. This narrow conceptualisation of generalizability represents a significant gap in existing validation frameworks for anatomical generative models, as it fails to capture the broader notion of generalisation in probabilistic generative settings. Mathematically, it is formulated as:

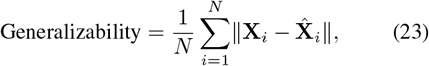

where *N* is the total number of shapes in the dataset, *X*_*i*_ is the ground truth shape, 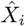 is the shape reconstructed using the model, ∥·∥ is the norm used to calculate the reconstruction error.

The evaluation of generalizability employs five main categories of metrics: Point-to-Point Distance Metrics, Set-Based Distance Metrics, Volumetric Overlap Metrics, Directional Similarity Metrics, and Anatomy-Specific Metrics.

#### 5.3.1 Point-based Distance Metrics

Point-based distance metrics measure direct distances between corresponding points in two datasets.

The **Euclidean Distance (ED)** is used to calculate the surface similarity of the heart [Dou et al., 2023, 2025, Kalaie et al., 2024].

The Symmetric Euclidean Distance (ED^*^) metric is typically used in cases where there is no point correspondence, requiring bidirectional measurements to properly capture the distance between sets:

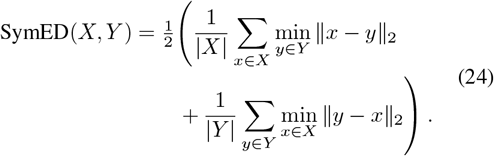

where *X* = (*x*_1_, *x*_2_, …, *x*_*n*_) are the ground truth set and *Y* = (*y*_1_, *y*_2_, …, *y*_*n*_) are the reconstructed set. Kalaie et al. [2024] applied it to provide a bidirectional measurement of the heart mesh sets.

The Root Mean Square Error (RMSE) represents the square root of the average of squared differences between the ground truth and the reconstructed shapes. It is formulated as:

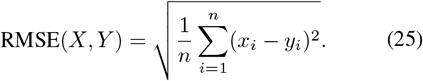

It is used to quantify the overall difference between reconstructed and ground truth occlusal surfaces of teeth [Hwang et al., 2018, Tian et al., 2022, Yuan et al., 2020] and heart shapes [Beetz et al., 2022b].

#### 5.3.2 Set-Based Distance Metrics

Set-based distance metrics evaluate distances between the ground truth and the reconstructed point sets without requiring explicit point correspondences.

The CD formulated as Eq. 33 calculates bidirectional average nearest neighbour distances between point sets of heart chambers [Seale et al., 2024, Beetz et al., 2022b, Peng et al., 2024], aortic root [Gasparovici and Serban, 2024], and blood vessels [Sinha and Hamarneh, 2024].

The Hausdorff Distance (HD) is a metric that measures how far two subsets of a metric space are from each other, focusing on the maximum deviation between shapes. The formulation of HD can be written as:

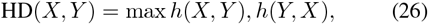

where *h*(*X, Y*) = *max*_*x*∈*X*_*min*_*y*∈*Y*_|| *x* − *y*|| is the directed HD from *X* to *Y*. It has been applied in cardiac shapes [Kalaie et al., 2024, Qiao et al., 2025, Xia et al., 2022, Sørensen et al., 2024] and jaw bone shapes [Abdi et al., 2019].

The Authenticity metric is a part of the 3-dimensional evaluation framework proposed by Alaa et al. [2022], which measures the generalisation capability of a generative model - specifically, it quantifies the extent to which the model generates new samples rather than copying training data. The authors formulate the generative distribution as a mixture:

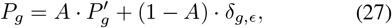

where *A* is the Authenticity score (between 0 and 1) which represents the probability that a synthetic sample was truly generated, *P′*_*g*_ is the generative distribution conditioned on samples not being copied, and *δ*_*g,ϵ*_ is a noisy distribution over training data. A synthetic sample is deemed “unauthentic” if it is closer to a specific training sample than any other training sample is to that same training sample. Thus, the Authenticity score can be expressed as:

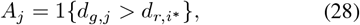

where *A*_*j*_ is the authenticity score for a specific synthetic sample *j*, 1 {*condition*} is an indicator function that equals 1 when the condition inside is true, and 0 when it is false, *d*_*g,j*_ is the distance between synthetic sample *j* and its closest sample in the training dataset, and 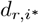 is the distance between that closest training sample and its nearest neighbour in the training set.

Although it has not been applied in the medical domain yet, it is still worth discussing as a potentially valuable evaluation metric.

#### 5.3.3 Volumetric Overlap Metrics

Volumetric overlap metrics quantify the spatial overlap between volumes.

Dice score, also known as F1 score, is represented as:

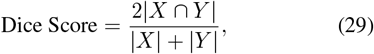

where |*X* ∩ *Y* | is the size of the intersection between sets *X* and *Y*, and | · | represents the total number of voxels in the binary maps.

Intersection over Union (IoU), also known as the Jaccard Index, is calculated as:

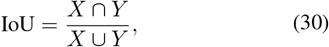

and *X* ∪ *Y* represents the union of the two sets. Both Dice Score and IoU are used to evaluate the similarity between the generated dental crown and technicians’ designs [Hwang et al., 2018], as well as between the reconstructed jaws and the ground truth segmentations [Abdi et al., 2019].

#### 5.3.4 Directional Similarity Metrics

Directional similarity metrics assess the angular alignment of features while being magnitude-invariant.

Cosine Similarity (CosSim) measures the angular similarity between feature vectors:

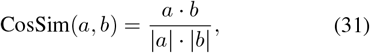

where *a* and *b* are vectors in a multi-dimensional space, *a· b* is the dot product of vectors *a* and *b*, and | · | is the magnitudes [Feldman et al., 2023].

#### 5.3.5 Anatomy-Specific Metrics

Anatomy-specific metrics are designed to evaluate particular anatomical characteristics.

The surface error (Error-surf) normalises nodal distance errors by the distance to the cross-sectional centroid, while the cortical thickness error (Error-thck) measures the ratio between thickness reconstruction error and ground truth of the rib [Huang et al., 2024].

These metrics collectively provide a comprehensive framework for evaluating a model’s generalisation capabilities. The selection of specific metrics often depends on the particular application requirements and the nature of the data being generated.

### 5.4 Diversity

Diversity evaluates whether the generated samples sufficiently represent the full range of features and patterns in the real data [Alaa et al., 2022].

#### 5.4.1 Activity

Activity is a quantitative metric used to evaluate the effectiveness of latent space representations in synthetic data generation models. This metric assesses how meaningfully each dimension of the latent space captures and encodes information from the input data, providing insights into the model’s ability to learn and represent important data characteristics. It is calculated as:

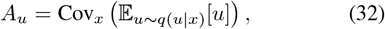

where *A*_*u*_ is the activity statistic for the latent dimension *u, q*(*u*|*x*) is the approximate posterior distribution for the latent variable *u* given the input *x*, 𝔼_*u*∼*q*(*u*|*x*)_[*u*] is the mean of *u* under the posterior *q*(*u* | *x*), and Cov_*x*_ is the covariance across the dataset. As demonstrated [Dou et al., 2023], activity measurement can be used to evaluate the diversity of generated samples by analysing how actively different latent dimensions participate in the generation process. This understanding of latent space is crucial for developing more efficient and effective synthetic VP generation systems in ISTs.

#### 5.4.2 Coverage Score

The COV assesses how well a generative model captures the full range of geometric variations present in the real underlying population. The Coverage Score is calculated by determining the percentage of shapes in the reference set that are matched by at least one generated sample, using a specified distance threshold for matching. The distance can be either Chamfer Distance (CD) or EMD as formulated in Eq. 15. CD can be represented as:

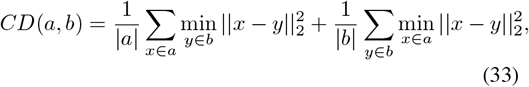

where *a, b* are point sets representing two shapes, |*a*|, |*b*| are the number of point in sets *a* and *b* respectively, *x, y* are individual points from sets *a* and *b*, and ||*x* − *y*||_2_ are Euclidean distance between points *x* and *y*. Thus, COV can be expressed as:

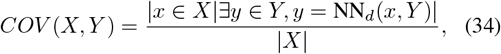

where *X* is the real set, *Y* is the generated set, and NN_*d*_ is the nearest neighbour function using either CD or EMD. A higher Coverage Score indicates that the generated population better spans the space of possible shape variations observed in real anatomical structures.

Peng et al. [2024] calculated the COV for the generations of multiple organs. Other applications in dental [Zhang et al., 2024a], vascular [Sinha and Hamarneh, 2024], and cardiac area [Peng et al., 2024] also calculated the COV to validate the diversity of their models when creating VPs for ISTs.

#### 5.4.3 β-Recall

The *β*-Recall metric is also a part of Alaa et al. [2022]’s framework that measures the diversity of generated samples - specifically, it quantifies the fraction of real samples that are covered by the generative distribution. It is defined as the probability that a real sample resides within the *β*-support of the generative distribution:

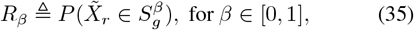

Wherev 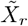 represents an embedded real sample (for the embedding details, please refer to Alaa et al. [2022]), and 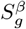 is the *β*-support of the generative distribution (which is the minimum volume subset of the support that contains a probability mass of *β*).

This framework has yet to be applied in any VP generation tasks for model evaluation, but it highlights the potential direction of developing a comprehensive framework to evaluate the generative models.

## 6 Meta Analysis

Our meta-analysis examines the performance of deep generative models of anatomy across four key aspects: static cardiac anatomy, dynamic cardiac anatomy, cardiac function, and other anatomies. The analysis encompasses various evaluation metrics and model architectures and excludes qualitative evaluations such as visual comparison, and each value of metrics in the tables is directly from the original paper. Ideally, the performance comparison should cover the four aspects of evaluation, and the values should be obtained by implementing the methods on the same dataset, yet it is not practical to achieve since the evaluations from each paper do not follow the same standards.

The number of publications using the same metric to evaluate a certain aspect is limited. Thus, we only compared the reconstruction error in the Dice score at the subgroup level. For the other metrics, we list the values for each individual method to compare their performance in one of the four aspects.

### 6.1 Static Cardiac Anatomy Performance

This part examines various approaches for static cardiac anatomy generation, evaluating their performance through both sample-level and population-level metrics, as shown in Table 16. At the sample level, reconstruction error and specificity error using Euclidean distance measurements provide insights into individual method performance, while population-level metrics, including MMD, COV, and clinical acceptance rates, offer broader evaluative perspectives. As mentioned in the evaluation metrics section, there exists a trade-off between generalisation and fidelity. We compared the performance of individual methods in terms of both reconstruction error and specificity error using ED. Analysis of reconstruction error using Dice scores reveals varying performance across model architectures, as shown in Fig. 11, with diffusion-based models demonstrating particularly strong results, achieving up to 97.80 ± 1.60% accuracy for Left Ventricle (LV) generation. Notably, VAE-based methods show consistent performance across multiple cardiac sub-anatomies, though with generally lower accuracy ranges of 70-90%. However, there still exists an imbalanced distribution of evaluation metrics across different model types, and anatomical structures present a challenge in making definitive comparative assessments. Population-level metrics, while less frequently reported, indicate promising results where available, with clinical acceptance rates reaching 99.90% for specific applications. Our analysis reveals significant methodological gaps, particularly in the comprehensive evaluation of population-level metrics across different anatomical structures. This limitation is particularly evident in the evaluation of whole heart structures, where only a subset of methods report comprehensive metrics. The observed trade-off between generalisation and fidelity underscores the need for standardised evaluation protocols that incorporate both sample-level and population-level metrics for a more comprehensive comparison of VP generation methods.

**Table 1:** Unconditional VAE methods. **Abbreviation:** Ao: Aorta, cMRI: Cardiac Magnetic Resonance Imaging, CT: Computed Tomography, D: Dynamic, ECG: Electrocardiogram, HCM: Hypertrophic Cardiomyopathy, LA: Left Atrium, LV: Left Ventricle, MRA: Magnetic Resonance Angiography, MRI: Magnetic Resonance Imaging, NA: Not applicable, NS: Not specified, RA: Right Atrium, RV: Right Ventricle, S: Static, US: Ultrasound.

| Ref | Anatomy | Model | Input | Output | D/S | Image Modality | Dataset Size | Synthetic Size | Health Condition |
| --- | --- | --- | --- | --- | --- | --- | --- | --- | --- |
| Danu et al. [2019] | Blood Vessel | VAE | 3D Binary Map | 3D Binary Map | S | NA | NS | 10000 | NS |
| HiREC-CVAE Cerrolaza et al. [2018] | Skull | cVAE | 2D Image | Binary Map | S | US | 72 | NA | NS |
| Biffi et al. [2019] | LV myo | cVAE | 2D Image | Binary Map | S | MRI | 1912 | NA | Healthy |
| Dou et al. [2025] | LV, RV, LA, RA, Ao | VAE | Surface Mesh | Surface Mesh | S | MRI | 2360 | 2000 | NS |
| Kalaie et al. [2024] | LV | VAE | Surface Mesh | Surface Mesh | S | MRI | 1000 | NA | NS |
| Biffi et al. [2018] | LV myo | VAE | 3D Binary Map | Binary Map, Class | S | MRI | 1365 | NA | HCM, Healthy |
| Beetz et al. [2022b] | LV endo/epi, RV | β-VAE | Point Cloud, ECG | Point Cloud, ECG | S | MRI | 539 | NA | Healthy |
| Abdi et al. [2019] | Mandible | cVAE | 3D Binary Map | 3D Binary Map | S | CT | 48 | NA | Defected Mandible |
| VesselVAE Feldman et al. [2023] | Blood Vessel | RVAE | Surface Mesh | Surface Mesh | S | MRA | 700 | NA | Healthy |
| Seale et al. [2024] | LV, RV | β-VAE | Point Cloud | Point Cloud | S | MRI | 539 | NA | Healthy |
| HybridVNet Gaggion et al. [2025] | LV, RV, LA, RA, Ao | Hybrid VNet | 2D/3D Image | Mesh | S | MRI | 4525 | NA | NS |

**Table 2:** Unconditional GAN methods. **Abbreviation:** AAA: Ascending Aortic Aneurysm, AD: Alzheimer’s Disease, Ao: Aorta, CT: Computed Tomography, D: Dynamic, Fluorescence Microscopy, GCN: Graph Convolutional Network, LSTM: Long Short-Term Memory, LV: Left Ventricle, MRI: Magnetic Resonance Imaging, NA: Not applicable, NS: Not specified, PN: PointNet, S: Static, SP-GAN: Sphere-guided Generative Adversarial Network.

| Ref | Anatomy | Models | Input | Output | D/S | Modality | Dataset Size | Synthetic Size | Health Condition |
| --- | --- | --- | --- | --- | --- | --- | --- | --- | --- |
| Danu et al. [2019] | Blood Vessel | GAN | 3D Binary Map | 3D Binary Map | S | NA | NS | 10000 | NS |
| HSPN [Hu et al., 2024a] | Brain | GAN, GCN | 2D Image | Point Cloud | S | MRI | 1040 | NA | AD |
| Chafi et al. [2024] | Dental Crown | SP-GAN | Point Cloud | Point Cloud | S | 3D Scan | 129 | NA | NS |
| Rezaei and Ahmadi [2023] | Lung Tumor | GAN, LSTM | 2D Binary Map | Surface Mesh | S | CT | 888 | NA | Nodule, No-nodule |
| Hu et al. [2021] | Brain | GAN | 2D Image | Point Cloud | S | MRI | 1040 | NA | AD, Healthy |
| Romero et al. [2021] | Ao | GAN, PCA | Surface Mesh | Surface Mesh | S | CT | 26 | 3000 | AAA |
| Gu and Gao [2023] | Lung Tumor | LSTM, GAN, ResNet | 2D Image | 3D Binary Map | S | CT | 888 | NA | Nodule, No-nodule |
| CranGAN [Sulakhe et al., 2022] | Skull | GAN, PN | Point Cloud | Point Cloud | S | CT | 210 | NA | NS |
| GAN-LSTM-3D [Hong et al., 2023] | Lung Tumor | GAN, LSTM | 2D Image | 3D Binary Map | S | CT | 888 | NA | Nodule, No-nodule |
| Wiesner et al. [2019] | Cell | GAN | 3D Binary Map | 3D Binary Map | S | FM | 385 | 770 | NS |
| Hu et al. [2025] | LV | GAN | 2D Image | Point Cloud | S | MRI | 1030 | NA | AD, Healthy |

**Table 3:** Unconditional Normalising Flow methods. **Abbreviation:** D: Dynamic, LV: Left Ventricle, MRI: Magnetic Resonance Imaging, NA: Not applicable, RV: Right Ventricle, S: Static

| Ref | Anatomy | Models | Input | Output | D/S | Modality | Dataset Size | Synthetic Size | Health Condition |
| --- | --- | --- | --- | --- | --- | --- | --- | --- | --- |
| Peng et al. [2024] | LV<br>endo/epi,<br>RV | Normaling<br>Flow,<br>Trans-<br>former | Point<br>Cloud | Point<br>Cloud | S | MRI | 994 | NA | Healthy |

**Table 4:** Unconditional Diffusion methods. **Abbreviation:** ACS: Abnormal Chamber Sizes, CCSD: Cardiac Conduction System Disorders, cMRI: Cardiac Magnetic Resonance Imaging, CT: Computed Tomography, CTA: Computed Tomography Angiography, D: Dynamic, LBBB: Left Bundle Branch Block, MRA: Magnetic Resonance Angiography, MRI: Magnetic Resonance Imaging, NA: Not applicable, NS: Not specified, S: Static, SD: Septal Defects, Trans: Transformer, VTV: Ventricular Trabeculae Variations.

| Ref | Anatomy | Models | Input | Output | D/S | Image Modality | Dataset Size | Synthetic Size | Health Condition |
| --- | --- | --- | --- | --- | --- | --- | --- | --- | --- |
| TrinD<br>[Sinha and Hamarneh, 2024] | Blood Vessel | DDM, Trans | Volumetric Mesh | Volumetric Mesh | S | CT, MRI, MRA, CTA | 186 | 120 | NS |
| DMCVR<br>[He et al., 2023] | LV, RV | DDIM | 2D+t Image | 3D Binary Map | S | cMRI | 1008 | NA | Healthy |
| Jayakumar et al. [2023] | LV | DDIM | 2D Image | 3D Binary Map | S | MRI | 215 | NA | CCSD, LBBB |
| Kadry et al. [2024b] | Coronary Artery | LDM | 3D Binary Map | 3D Binary Map | S | CT | 808 | NA | Calcification, LBBB |
| Kadry et al. [2024a] | Ao, LV, RV, LA, RA | DDIM | 3D Binary Map | 3D Binary Map | S | CT | 512 | NA | SD, ACS, VTV |

**Table 5:** Unconditional DeepSSM methods. **Abbreviation:** CT: Computed Tomography, D: Dynamic, DeepSDF: Deep Signed Distance Function, DeepSSM: Deep Learning-based Statistical Shape Models, LS: Laser Scanning, NA: Not applicable, NS: Not specified, S: Static.

| Ref | Anatomy | Models | Input | Output | D/S | Modality | Dataset Size | Synthetic Size | Health Condition |
| --- | --- | --- | --- | --- | --- | --- | --- | --- | --- |
| Nauwelaers et al. [2021] | Palatal | DeepSSM | Surface Mesh | Surface Mesh | S | LS | 2648 | NA | NS |
| Gasparovici and Serban [2024] | Aortic root | DeepSDF | Surface Mesh | Surface Mesh | S | CT | 97 | NA | NS |

**Table 6:** Conditional VAE methods. **Abbreviation:** AD: Alzheimer’s Disease, cMRI: Cardiac Magnetic Resonance Imaging, CT: Computed Tomography, D: Dynamic, HC: Hippocampus, HCM: Hypertrophic Cardiomyopathy, LSTM: Long Short-Term Memory, MRI: Magnetic Resonance Imaging, Myo: Myocardium, NA: Not applicable, NS: Not specified, PMI: Previous Myocardial Infarction, S: Static, Trans: Transformer.

| Ref | Anatomy | Models | Conditioning | Input | Output | D/S | Modality | Dataset Size | Synthetic Size | Health Condition |
| --- | --- | --- | --- | --- | --- | --- | --- | --- | --- | --- |
| Huang et al. [2024] | Rib | cVAE | Sex, Rib, Age, Height | 3D Binary Map | 3D Binary Map | S | CT | 3193 | NA | NS |
| Qiao et al. [2025] | LV, RV, Myo | VAE, Trans | Age, Sex, Weight, Height | Surface Mesh | Surface Mesh | D | cMRI | 38309 | NA | Healthy |
| CHearT Qiao et al. [2024] | LV, RV, Myo | β-VAE, LSTM | Age, Sex, Clinical | 3D+t | 3D+t | D | cMRI | 1383 | NA | Healthy |
| Beetz et al. [2023] | LV endo/epi, RV | β-VAE | Sex, Phase | Point Cloud | Point Cloud | S | cMRI | 1000 | NA | Healthy |
| Biffi et al. [2020] | LV, HC | LVAE | Clinical | 3D Binary Map | 3D Binary Map | S | MRI | 1613 | NA | HCM, Healthy, AD |
| Beetz et al. [2022d] | LV | β-VAE | Phase | Surface Mesh | Surface Mesh | S | cMRI | 1021 | NA | PMI |
| Foti et al. [2020] | Liver | VAE | Part of Liver | Point Cloud, Mesh | Surface Mesh | S | CT | 50 | 4500 | NS |

**Table 7:** Conditional GAN methods. **Abbreviation:** CTA: Computed Tomography Angiography, D: Dynamic, DCPR-GAN: Dental Crown Perception Refinement Generative Adversarial Network, GP-GAN: Growth Prediction Generative Adversarial Network, IS: Intraoral Scanner, L/HGG: Low/High-Grade Glioma, MRI: Magnetic Resonance Imaging, NA: Not applicable, NS: Not specified, RV: Right Ventricle, S: Static, T1: T1-weighted.

| Ref | Anatomy | Models | Conditioning | Input | Output | D/S | Modality | Dataset Size | Synthetic Size | Health Condition |
| --- | --- | --- | --- | --- | --- | --- | --- | --- | --- | --- |
| Wolterink et al. [2018] | Coronary Artery | WGAN | Vessel Characteristics | Central Line | Central Line | S | CTA | 4412 | NA | NS |
| DCPR-GAN [Tian et al., 2022] | Dental Crown | GAN | Tooth Parameters | 2D Image | Surface Mesh | S | 3D Scan | 780 | NA | NS |
| GP-GAN [Elazab et al., 2020] | Brain Tumor | CGAN | T1 Features | 3D Binary Map | 3D Binary Map | D | MRI | 6375 | NS | L/HGG |
| Hwang et al. [2018] | Dental Crown | CGAN | NA | 2D Image | Surface Mesh | S | 3D Scan | 3313 | NA | NS |
| Yuan et al. [2020] | Dental Occlusal | CGAN | NA | 2D Image | Surface Mesh | S | IS | 600 | NA | Healthy |
| Qiao et al. [2022] | LV, RV | CycleGAN | Age | 3D Binary Map | 3D Binary Map | D/S | MRI | 12600 | NA | Cardiovascular Disease |

**Table 8:** Flow-based conditional methods. **Abbreviation:** CHD: Congenital Heart Disease, CT: Computed Tomography, cVAE: Conditional Variational Autoencoder, cVAE-NF: Conditional Variational Autoencoder with Normalising Flow, D: Dynamic, LV: Left Ventricle, MRI: Magnetic Resonance Imaging, NA: Not applicable, NF: Normalising Flow, NS: Not specified, S: Static, SDF: Signed Distance Function.

| Ref | Anatomy | Models | Conditioning | Input | Output | D/S | Modality | Dataset Size | Synthetic Size | Health Condition |
| --- | --- | --- | --- | --- | --- | --- | --- | --- | --- | --- |
| cVAE-NF Dou et al. [2023] | LV | cVAE, NF | Clinical | Surface Mesh | Surface Mesh | S | MRI | 2360 | NA | NS |
| Kong et al. [2024] | Whole Heart | NF | CHD Type | SDF | SDF | S | CT | 110 | NA | CHD |

**Table 9:**
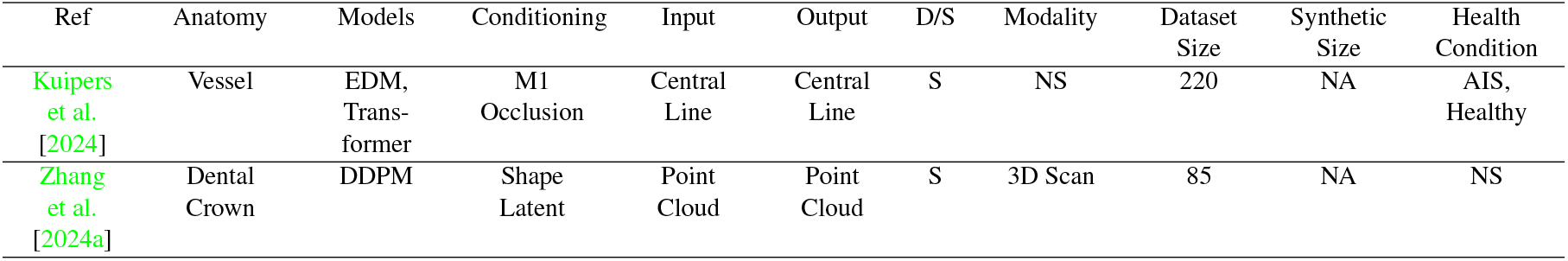
Conditional Diffusion methods. **Abbreviation:** AIS: Acute Ischemic Stroke, D: Dynamic, DDPM: Denoising Diffusion Probabilistic Models, EDM: Elucidated Diffusion Model, M1: Middle Cerebral Artery M1 Segment, NA: Not applicable, NS: Not specified, S: Static.

| Ref | Anatomy | Models | Conditioning | Input | Output | D/S | Modality | Dataset Size | Synthetic Size | Health Condition |
| --- | --- | --- | --- | --- | --- | --- | --- | --- | --- | --- |
| Kuipers et al. [2024] | Vessel | EDM, Trans-former | M1 Occlusion | Central Line | Central Line | S | NS | 220 | NA | AIS, Healthy |
| Zhang et al. [2024a] | Dental Crown | DDPM | Shape Latent | Point Cloud | Point Cloud | S | 3D Scan | 85 | NA | NS |

**Table 10:**
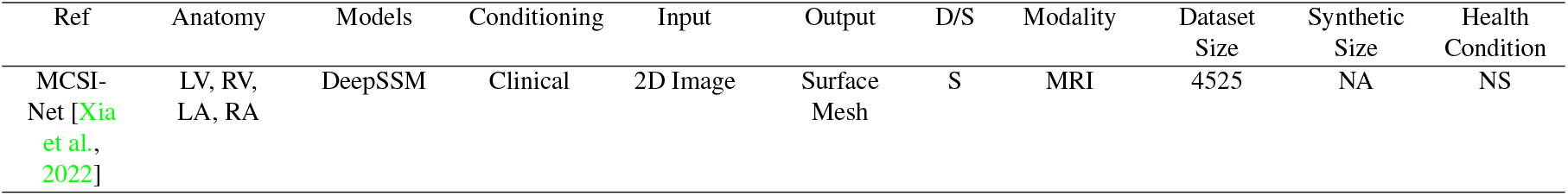
Conditional DeepSSM methods. **Abbreviation:** D: Dynamic, LA: Left Atrium, LV: Left Ventricle, MCSI-Net: Multi-Cue Shape Inference Network, MRI: Magnetic Resonance Imaging, NA: Not applicable, NS: Not specified, RA: Right Atrium, RV: Right Ventricle, S: Static.

**Table 11:**
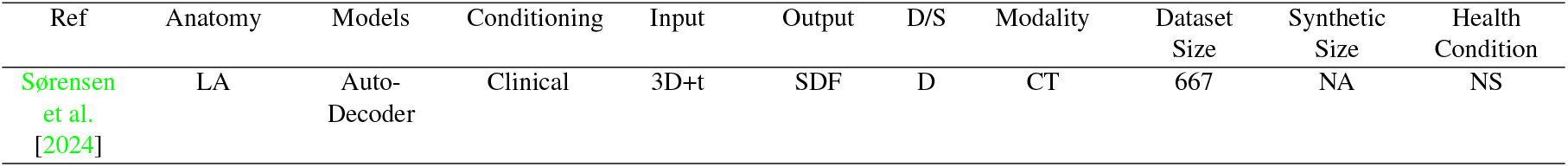
Other conditional methods. **Abbreviation:** CT: Computed Tomography, D: Dynamic, LA: Left Atrium, NA: Not applicable, NS: Not specified, S: Static, SDF: Signed Distance Function

| Ref | Anatomy | Models | Conditioning | Input | Output | D/S | Modality | Dataset Size | Synthetic Size | Health Condition |
| --- | --- | --- | --- | --- | --- | --- | --- | --- | --- | --- |
| Sørensen et al. [2024] | LA | Auto-Decoder | Clinical | 3D+t | SDF | D | CT | 667 | NA | NS |

**Table 12:** Evaluation Methods for Fidelity Assessment of Generated Synthetic Populations.

| Metric Type | Evaluation Study |  | Ref |
| --- | --- | --- | --- |
| Shape-based | 1st Nearest Neighbour Accuracy |  | Peng et al. [2024], Sinha and Hamarneh [2024] |
|  | Divergence | Earth Mover’s Distance | Qiao et al. [2025, 2024, 2022] |
|  |  | Jensen-Shannon Divergence | Zhang et al. [2024a] |
|  |  | Kullback-Leibler Divergence | Huang et al. [2024], Qiao et al. [2025, 2024, 2022] |
|  | Maximum Mean Discrepancy |  | Beetz et al. [2022b] |
|  | Minimum Matching Distance | Chamfer Distance | Zhang et al. [2024a], Sinha and Hamarneh [2024], Peng et al. [2024], Kadry et al. [2024a] |
|  |  | Earth Mover’s Distance | Zhang et al. [2024a], Sinha and Hamarneh [2024], Peng et al. [2024], Kadry et al. [2024a] |
|  | Specificity |  | Dou et al. [2023, 2025], Kalaie et al. [2024], Kuipers et al. [2024] |
|  | Statistical Test | Kolmogorov-Smirnov Test | Wiesner et al. [2019], Kuipers et al. [2024] |
|  |  | Mann-Whitney-Wilcoxon Test | Romero et al. [2021] |
| Feature-based | Anthropometric Analysis |  | Qiao et al. [2025], Dou et al. [2023], Wolterink et al. [2018], Qiao et al. [2024], Romero et al. [2021], Kuipers et al. [2024], Hwang et al. [2018], Sørensen et al. [2024], Feldman et al. [2023], Sinha and Hamarneh [2024], Huang et al. [2024], Dou et al. [2025], Tian et al. [2022], Beetz et al. [2022b], Wiesner et al. [2019] |
|  | Interpolation Study |  | Wolterink et al. [2018], Beetz et al. [2022b], Kong et al. [2024] |

**Table 13:** Evaluation Methods for Utility Aspect of Generated Synthetic Populations.

| 1st Level Metric | Ref |
| --- | --- |
| Clinical Acceptance Rate | Dou et al. [2025],<br>Kalaie et al. [2024],<br>Romero et al. [2021] |
| Clinician Validation | Abdi et al. [2019],<br>Hwang et al. [2018] |
| Downstream Classification | Romero et al. [2021],<br>Beetz et al. [2022b],<br>Qiao et al. [2025] |
| Downstream Prediction | Huang et al. [2024] |

**Table 14:** Evaluation Methods for Generalizability Assessment of Generated Synthetic Populations.

| 1st Level Metric | Metric Type | 2nd Level Metric | Ref |
| --- | --- | --- | --- |
| Reconstruction Error | Point-to-Point Distance Metrics | Euclidean Distance | Dou et al. [2023, 2025], Kalaie et al. [2024] |
|  |  | Symmetric Euclidean Distance | Kalaie et al. [2024] |
|  |  | Root Mean Square Error | Hwang et al. [2018], Tian et al. [2022], Yuan et al. [2020], Beetz et al. [2022b] |
|  |  | Peak Signal-to-Noise Ratio | Tian et al. [2022], Yuan et al. [2020] |
|  |  | Average Symmetric Surface Distance | Qiao et al. [2025, 2024] |
|  | Set-Based Distance Metrics | Chamfer Distance | Seale et al. [2024], Beetz et al. [2022b], Sinha and Hamarneh [2024], Gasparovici and Serban [2024], Peng et al. [2024] |
|  |  | Symmetric Chamfer Distance | Sørensen et al. [2024] |
|  |  | Hausdorff Distance | Kalaie et al. [2024], Qiao et al. [2025], Xia et al. [2022], Sørensen et al. [2024], Abdi et al. [2019] |
|  | Volumetric Overlap Metrics | Dice Score | Wiesner et al. [2019], Xia et al. [2022], Abdi et al. [2019], Dou et al. [2025], Qiao et al. [2022], Kong et al. [2024], He et al. [2023], Jayakumar et al. [2023] |
|  |  | Intersection over Union | Wiesner et al. [2019], Hwang et al. [2018], Kong et al. [2024] |
|  | Directional Similarity Metrics | Cosine Similarity | Feldman et al. [2023] |
|  | Anatomy-Specific Metrics | Surface Error | Huang et al. [2024] |
|  |  | Thickness Error | Huang et al. [2024] |

**Table 15:**
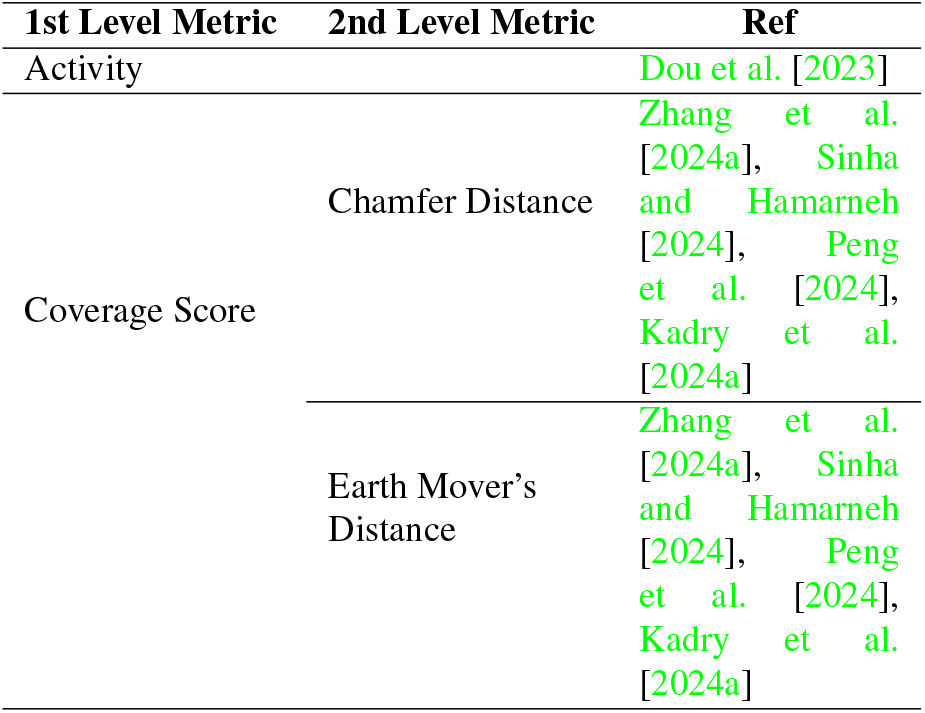
Virtual Population Diversity.

**Table 16:**
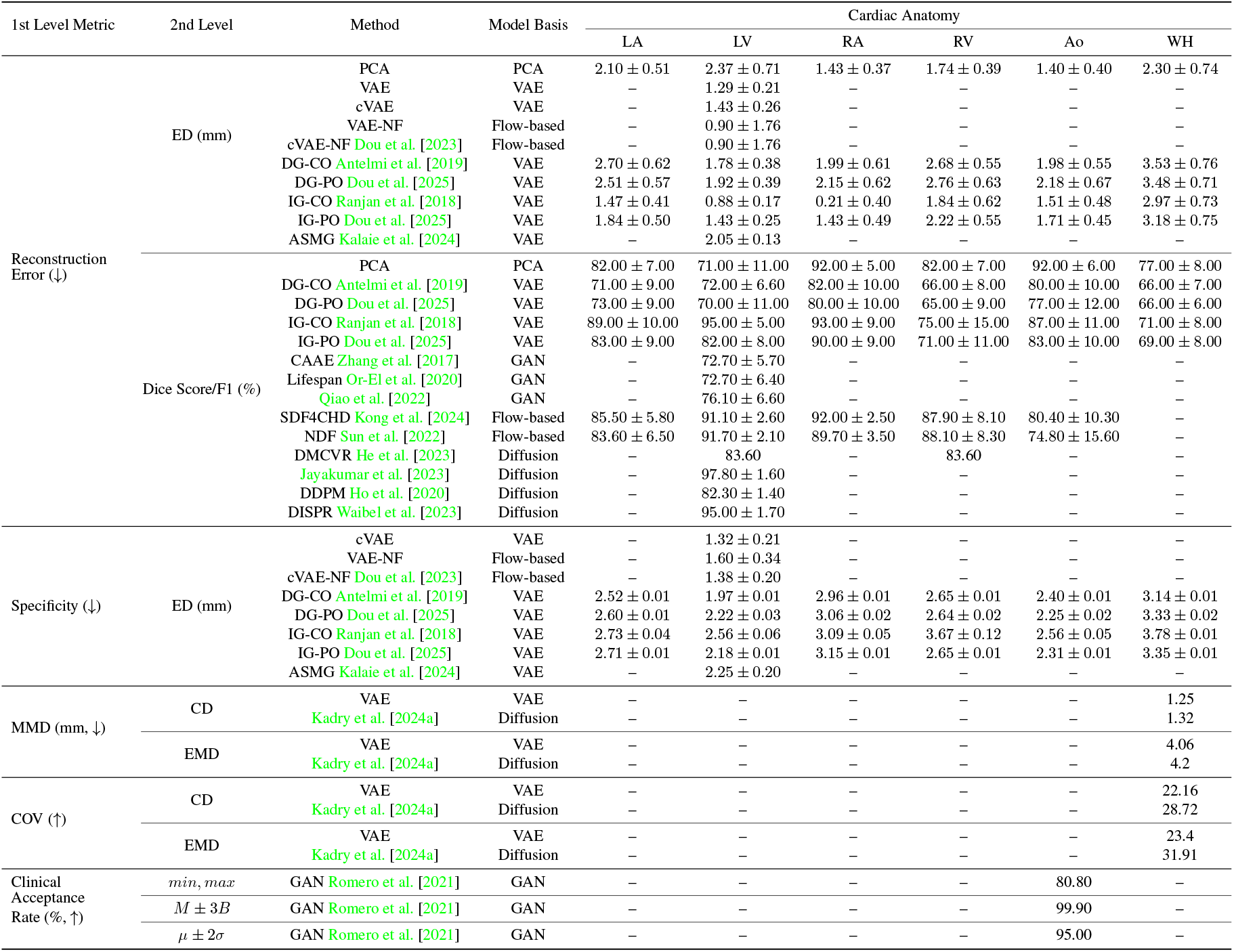
Performance Comparison of Methods on Static Cardiac Anatomy.

**Table 17:**
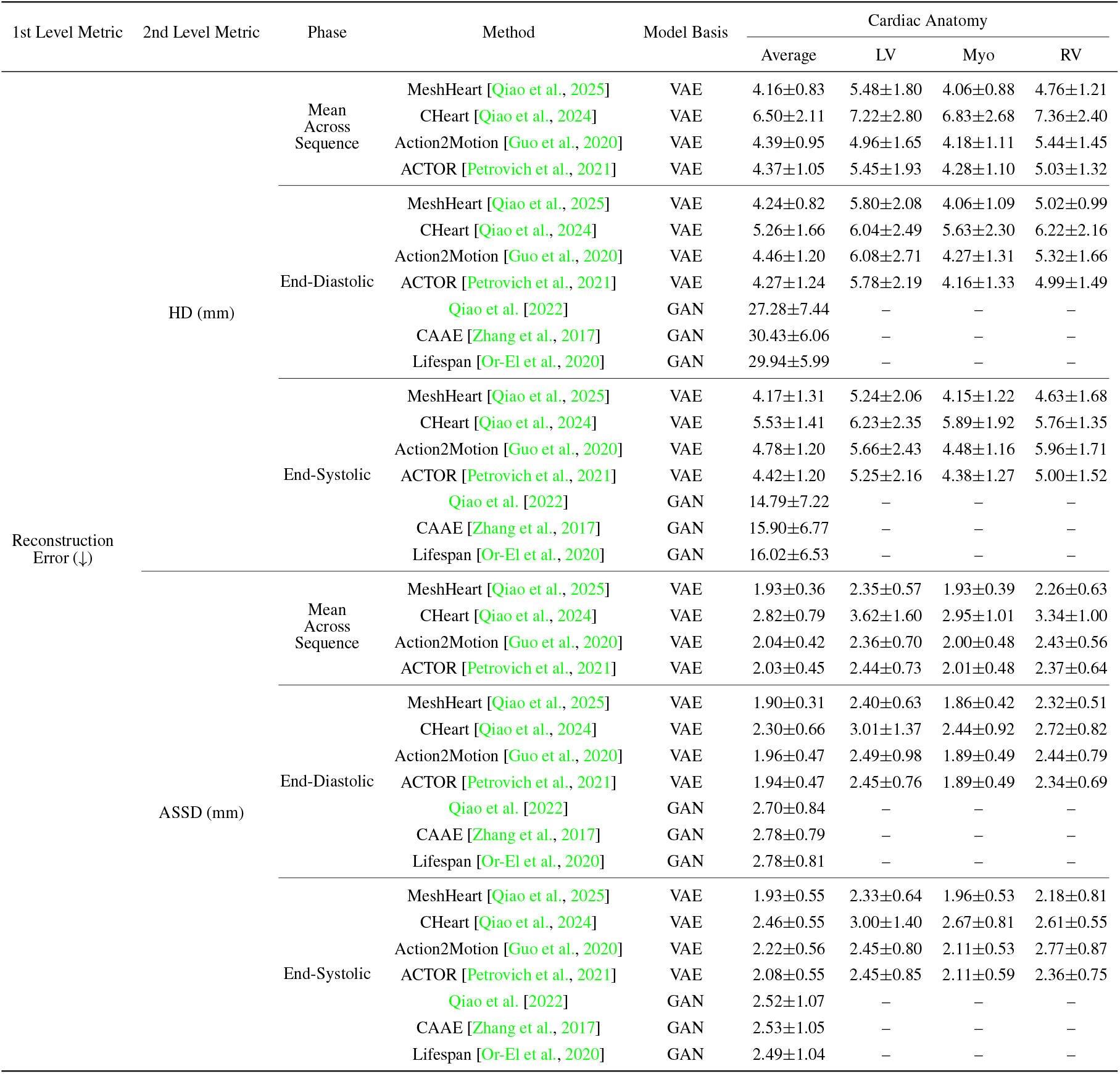
Performance Comparison of Methods on Dynamic Cardiac Anatomy.

**Table 18:**
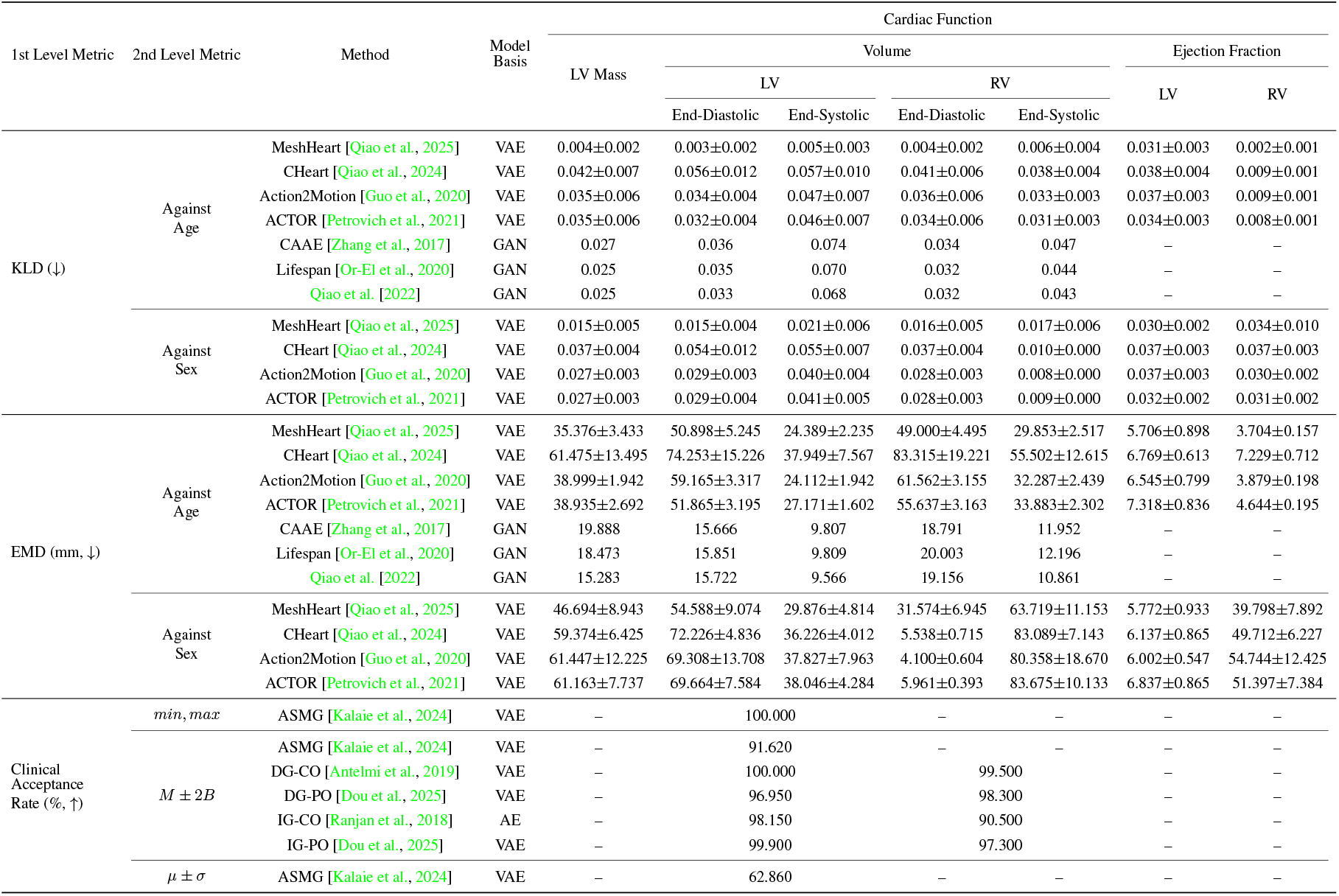
Performance Comparison of Methods on Cardiac Function.

**Table 19:** Performance Comparison of Methods on Other Anatomy.

| 1st Level Metric | 2nd Level Metric | Method | Model Basis | Other Anatomy |  |
| --- | --- | --- | --- | --- | --- |
|  |  |  |  | Vessel | Dental Crown |
| MMD (↓) | CD | TrIND [Sinha and Hamarneh, 2024] | Diffusion | 13.36 | – |
|  |  | PointFlow [Yang et al., 2019] | NF | – | 2.86 |
|  |  | DPM [Luo and Hu, 2021] | Diffusion | – | 2.46 |
|  |  | Zhang et al. [2024a] | Diffusion | – | 2.41 |
|  | EMD | PointFlow [Yang et al., 2019] | NF | – | 1.82 |
|  |  | DPM [Luo and Hu, 2021] | Diffusion | – | 1.68 |
|  |  | Zhang et al. [2024a] | Diffusion | – | 1.63 |
| COV (↑) | CD | TrIND [Sinha and Hamarneh, 2024] | Diffusion | 0.46 | – |
|  |  | PointFlow [Yang et al., 2019] | NF | – | 0.30 |
|  |  | DPM [Luo and Hu, 2021] | Diffusion | – | 0.36 |
|  |  | Zhang et al. [2024a] | Diffusion | – | 0.3 |
|  | EMD | PointFlow [Yang et al., 2019] | NF | – | 0.36 |
|  |  | DPM [Luo and Hu, 2021] | Diffusion | – | 0.42 |
|  |  | Zhang et al. [2024a] | Diffusion | – | 0.44 |

**Table 20:** Challenges and Future Directions in Anatomical Geometry Generation.

| Category | Key Challenges | Future Directions |
| --- | --- | --- |
| **Generation Methods** |  |  |
| **Data-related** | • Multi-modal data integration<br>• Dynamic generation<br>• Multi-resolution consistency maintenance | • Multi-modal frameworks<br>• Motion generation models<br>• Cross-resolution and super-resolution training |
| **Geometry/Topology** | • Topological complexity handling<br>• Semantic controlled generation<br>• Fine-scale feature capture | • Topology-aware loss design and Topology-adaptive models<br>• Disentangled generation<br>• Attention-guided refinement |
| **Physiological Plausibility** | • Physiologically plausible VP generation | • Anatomical and anthropometric constraints |
| **Evaluation** |  |  |
| **Comprehensive Evaluation** | • Not covering all aspects of evaluation<br>• Non-standardised evaluation metrics | • A systematic evaluation workflow composed of both sample-level and population-level evaluation metrics that covers fidelity, utility, generalizability, and diversity |
| **Hierarchical Validation** | • Evaluation only focusing on a fixed scale |  |
| **Clinical Translation** | • ISTs’ applicability<br>• Clinical outcome correlation | • Simulation-based evaluation<br>• VP generation - Systematic Evaluation - ISTs validation loop |

**Figure 11.**
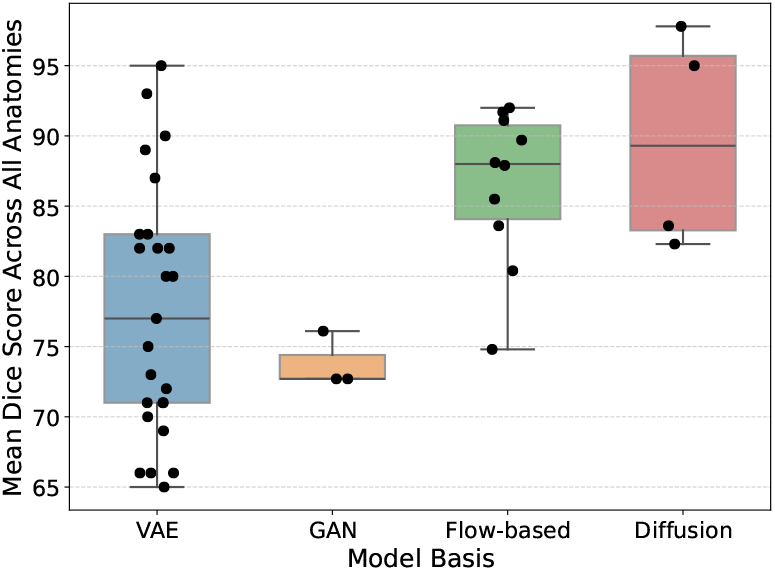
Dice Score between The Reconstructed Heart and The Ground truth by Model Basis

### 6.2 Dynamic Cardiac Anatomy Performance

We evaluated multiple methods using two primary error metrics: HD and ASSD across different cardiac cycle phases (End-Diastolic, End-Systolic, and mean sequence). A notable disparity emerged between VAE-based and GAN-based approaches, with VAE models demonstrating substantially superior performance. This performance gap can be attributed to the significant difference in training data volume, where VAE models benefited from a robust dataset of 38309 subjects compared to GAN models’ limited training set of 639 subjects. The HD metric revealed considerable variations, particularly evident in GAN models’ performance at the End-Diastolic phase (27.28-30.43mm) compared to VAE models (4.24-5.26mm). However, this dramatic difference highlighted HD’s susceptibility to outliers and poor-quality generated samples, underscoring the importance of metric selection in evaluation frameworks. In contrast, ASSD provided more stable measurements across all methods due to its averaging mechanism. Analysis of ASSD values demonstrated consistent superior performance of VAE-based methods, with Mesh-Heart Qiao et al. [2025] achieving the lowest error rates across all phases (mean sequence: 1.93 ± 0.36mm, End-Diastolic: 1.90 ± 0.31mm, End-Systolic: 1.93 ± 0.55mm), followed closely by ACTOR [Petrovich et al., 2021] and Action2Motion [Guo et al., 2020]. All VAE-based methods maintained ASSD values below 3mm, indicating reliable reconstruction quality, while GAN-based approaches showed marginally higher error rates ranging from 2.49mm to 2.78mm, though still within acceptable clinical thresholds.

We compared seven anatomical and functional parameters: LV Mass, LV and RV volumes at both End-Diastolic and End-Systolic phases, and LV and RV Ejection Fractions. The comparison encompassed seven methods using both KLD and EMD metrics against age and sex distributions. MeshHeart [Qiao et al., 2025] demonstrated superior performance in age-based distribution matching, achieving the lowest KLD scores across all metrics (ranging from 0.002 ± 0.001 to 0.031± 0.003), significantly outperforming other methods. In sex-based comparisons, MeshHeart and ACTOR [Petrovich et al., 2021] showed comparable performance, with KLD values generally below 0.03. Interestingly, while GAN-based approaches like Lifespan [Or-El et al., 2020] showed competitive KLD scores against age (ranging from 0.025 to 0.070), they lacked evaluation metrics for ejection fraction parameters. The EMD analysis revealed a different pattern, with GAN-based methods showing lower absolute distances (EMD values ranging from 9.566mm to 20.003mm) compared to VAE-based approaches, particularly in volume measurements. However, this apparent advantage should be interpreted cautiously, given their limited training dataset. Regarding clinical acceptance rates, methods employing different statistical criteria (min-max, *M* ± 2*B, µ* ± *σ*) showed varying levels of success. The DG-CO method achieved the highest acceptance rate (100% for LV and 99.5% for RV) under the *M* ± 2*B* criterion, while ASMG demonstrated perfect acceptance under min-max criteria but lower rates (62.86%) when evaluated using *µ* ± *σ*. These findings suggest that while VAE-based methods generally provide more robust and comprehensive cardiac function modelling, the choice of evaluation metric and acceptance criteria significantly impacts the assessment of generative model performance in cardiac imaging applications.

### 6.3 Other Anatomy

We evaluated the performance of various generative models on vessel and dental crown anatomies using MMD and COV metrics, each calculated with both CD and EMD where applicable. For vessel anatomy, only TrIND, a diffusion-based model, was evaluated, achieving an MMD-CD score of 13.36mm and a COV-CD of 0.46. In the dental crown domain, three methods were compared: PointFlow (based on Normalising Flows) [Yang et al., 2019], DPM [Luo and Hu, 2021], and Zhang et al.’s [Zhang et al., 2024a] approach (both diffusion-based). The diffusion-based methods demonstrated superior performance, with Zhang et al.’s method achieving the lowest MMD scores (CD: 2.41mm, EMD: 1.63mm), marginally outperforming DPM (CD: 2.46mm, EMD: 1.68mm) and more substantially surpassing PointFlow (CD: 2.86mm, EMD: 1.82mm). Interestingly, while diffusion models showed better performance in distance-based metrics, their advantage in coverage metrics was less pronounced. In COV-EMD, diffusion-based approaches (Zhang et al.: 0.44, DPM: 0.42) showed modest improvements over PointFlow (0.36), while COV-CD results were comparable across all methods (ranging from 0.30 to 0.36). These findings suggest that while diffusion-based models generally offer superior performance in geometric accuracy, the choice between different generative approaches may depend on specific application requirements and the relative importance of different evaluation metrics.

## 7 Discussion

In analysing the landscape of deep learning-based geometry generation methods for virtual populations, our comprehensive review reveals several significant patterns and challenges that merit careful consideration. These findings not only illuminate the current state of the field but also point toward promising future research directions that could advance the capabilities of virtual population generation. This discussion section examines two critical aspects of virtual population generation: the evolution and limitations of generative models and the current state of evaluation methodologies. Through this dual lens, we aim to provide insights that will help shape future research directions and address existing limitations in the field. By analysing both the modelling approaches and evaluation frameworks, we can better understand the interconnected challenges that must be overcome to advance the field of virtual population generation. Our analysis focuses particularly on how different architectural choices and evaluation metrics interact with the underlying anatomical complexity of various structures and how these interactions influence the quality and utility of generated populations. This perspective allows us to identify not only the technical challenges but also the broader implications for clinical applications and research utility.

### 7.1 Models

The strengths and limitations of models vary based on the choices of architectures. The prevalent use of VAE-based approaches in virtual population generation introduces certain limitations, particularly in capturing fine geometric details and complex topologies as shown in Danu et al. [2019], Feldman et al. [2023], as noted in studies like Biffi et al. [2019] and Beetz et al. [2022d]. Compared with virtual vessel population generation, which requires topology guidance and fine details, these limitations of VAE are more acceptable when generating a virtual heart population. In examining the methodological landscape of deep learning-based geometry generation, we observe distinct patterns in how different generative architectures align with specific anatomical structures, as shown in Fig. 12. Our analysis reveals that the choice of generative model is intrinsically linked to the topological complexity and anatomical constraints of the target structures, leading to varying degrees of success across different applications. This relationship between model architecture and anatomical complexity presents both opportunities and challenges that warrant careful consideration. This subsection examines how different generative architectures interact with various anatomical structures, highlighting both their strengths and limitations. By understanding these interactions, we can identify specific technical challenges that need to be addressed and propose potential directions for future research in the field of virtual population generation.

**Figure 12.**
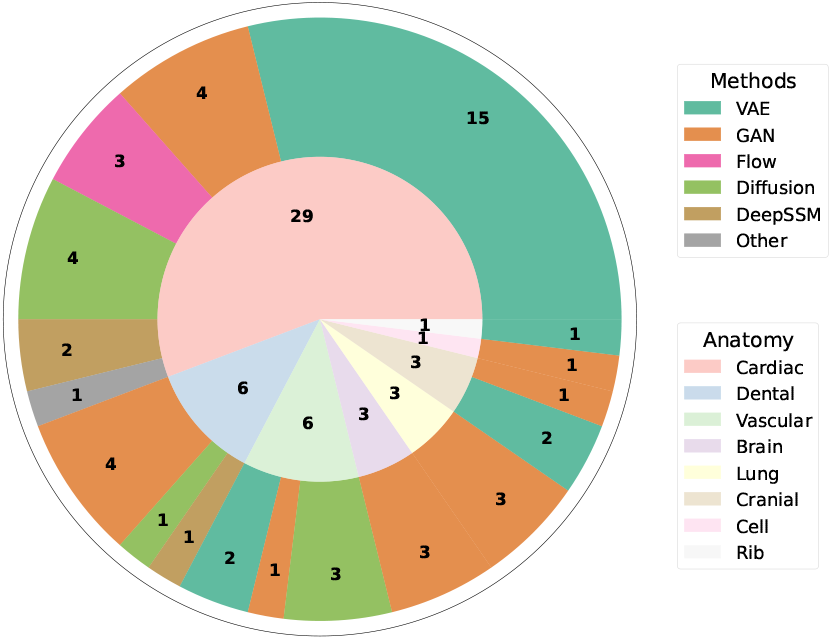
Methods Analysis of Methods on Model Basis and Anatomy. The numbers on the inner circle represent the number of publications about the specific anatomy. The numbers on the outer circle represent the number of publications using the specific method.

#### 7.1.1 Model Architecture Selection Based on Topological Complexity

When selecting an appropriate generative model for anatomical structure generation, researchers should consider the inherent topological complexity of the target anatomy as a primary factor. Our analysis suggests a general principle: structures with fixed topology and regular morphology are better suited for VAE-based approaches, while those with variable topology and intricate branching patterns benefit from diffusion models or specialised architectures. This topology-guided model selection can significantly improve generation quality and anatomical plausibility, potentially reducing the need for extensive post-processing and validation steps. We recommend considering both the topological invariants (features that must be preserved) and topological variants (features that may change across the population) when designing or selecting a generative architecture for specific anatomical applications.

The analysis reveals distinct methodological patterns across different anatomical structures. Among the 52 papers reviewed, cardiac geometries dominate the research landscape with 29 studies (55.8% of total), including whole heart (Beetz et al. [2022c,a,d, 2024], Biffi et al. [2019, 2018, 2020]), multi-chamber generation [Dou et al., 2023, 2025], congenital diseased heart generation [Kong et al., 2024], and recent advancements in dynamic cardiac generation [Qiao et al., 2025, 2024]. This cardiac-centric focus stems from two synergistic factors: (1) the clinical urgency of cardiovascular diseases accounting for 31% to 33% of global mortality [GBD 2019 Diseases and Injuries Collaborators, 2020, Timmis et al., 2022], and (2) the anatomical suitability of cardiac structures for deep learning approaches. Notably, VAE-based methods emerge as the predominant architecture in cardiac applications (15/29 cardiac studies), achieving particular success through their compatibility with the heart’s fixed topological characteristics. As demonstrated in anatomical studies [Padala et al., 2020, Hill et al., 2024, Whiteman et al., 2021], the cardiac system maintains a closed, single-connected domain topology with well-defined chamber relationships. This structural regularity enables smooth latent space embedding in VAEs, where continuous parameter variations correspond to physiologically plausible morphologies. The methodology aligns with established cardiac modelling paradigms that utilise population atlases for phenotype variation mapping [Young and Frangi, 2009, Lombaert et al., 2012, Bai et al., 2015], ranging from congenital defects [Abbott, 2006, Sophocleous et al., 2022] to acquired pathologies like obesity-related remodelling [Marciniak et al., 2021]. Such patterns can also be observed in other organs that share similar features in their structures, like the rib and cranial structures.

#### 7.1.2 Topological Considerations in Generative Models

The topological complexity of anatomical structures directly influences the effectiveness of different generative architectures [Uzunova et al., 2022], creating a fundamental relationship between anatomical characteristics and model performance. This relationship is particularly evident when comparing cardiac and vascular generation tasks, where the stark contrast in topological complexity necessitates different modelling approaches.

In contrast to cardiac structures, vascular systems exhibit unfixed and highly complex topological characteristics that pose fundamental challenges for geometric generation. Blood vessels demonstrate four key complexity dimensions: (1) tortuosity differences among normal and diseased vessels [Bullitt et al., 2003, 2005, Alilou et al., 2018]; (2) multi-scale spatial organization governed by Murray’s law for optimal flow distribution [Sherman, 1981]; (3) various diameters distribution in different vascular systems [Chen et al., 1994, Hanssen et al., 2022]; and (4) bifurcation angles [UYLINGS, 1977]. These discrete features conflict with VAE’s continuous latent space assumptions, as evidenced by early vessel generation attempts like Danu et al. [2019], Wolterink et al. [2018] that were limited to short straight segments with simplified bifurcations, suggesting a need for specialised architectures that can handle topological features like branching patterns, branching angles, and smoothness, etc.

This topological complexity directly correlates with the methodological landscape - vessel generation studies constitute only 6 of 52 reviewed papers [Wolterink et al., 2018, Feldman et al., 2023, Kuipers et al., 2024, Sinha and Hamarneh, 2024, Danu et al., 2019], showing no clear architectural dominance. However, recent advances in diffusion models demonstrate superior capability in handling vascular complexity, as Sinha and Hamarneh [2024] achieved in generating blood vessels with more complicated topology through stochastic progressive bifurcation. Similarly, Kuipers et al. [2024] utilised a hierarchical diffusion process to model cerebral vasculature with branch order preservation. These successes align with diffusion models’ theoretical strength in capturing discrete structures, making them particularly suitable for vascular generation tasks requiring explicit topology control, as evidenced in the topology optimisation area [Mazé and Ahmed, 2023, Zhu et al., 2024, Giannone et al., 2023]. The trend of using diffusion-based models for complex generation can also be easily observed from Fig. 3 in this review.

GAN-based approaches follow with 13 papers, dominating in the dental, brain, and lung [Hu et al., 2024a, 2021, 2025, Elazab et al., 2020] and lung [Rezaei and Ahmadi, 2023, Gu and Gao, 2023, Hong et al., 2023] applications. GAN architectures, despite their generative capabilities, have been predominantly applied to reconstruction tasks rather than direct population synthesis, with 8 of 14 GAN papers including Hu et al. [2024a], Gu and Gao [2023], and Sulakhe et al. [2022] focusing on potential benefit methods. This pattern is particularly evident in dental applications, where the primary goal is often to recover original tooth morphology rather than generate novel variations. This focus is driven by the complex constraints inherent in dental structures, including opposite tooth contact points, occlusal spatial relationships, and occlusal fingerprints - characteristics that must be precisely maintained to prevent dental complications and potential tooth damage. GANs demonstrate particular strengths in these reconstruction tasks for several reasons. First, their adversarial training mechanism naturally excels at preserving high-frequency details and sharp features, which is crucial for capturing the precise geometrical relationships in dental structures. Second, the discriminator network provides an implicit regularisation mechanism that helps maintain anatomical plausibility by learning to distinguish between realistic and unrealistic features. Third, GANs’ ability to learn hierarchical features allows them to capture both local details (such as cusps and ridges) and global structural relationships (like occlusal patterns) simultaneously. However, this same pattern may stem from challenges in controlling GAN outputs and ensuring anatomical validity in pure generation tasks. The success of GANs in reconstruction tasks, particularly for brain and lung structures, suggests the potential for adaptation to generation tasks through improved control mechanisms. This could potentially be achieved through the development of specialised architectural components or loss functions that explicitly encode anatomical constraints and relationships.

While DeepSSM and flow-based models demonstrate promising capabilities in shape analysis, their application to virtual population generation remains limited, particularly for complex anatomical structures like blood vessels. Current implementations of these methods, as reviewed in this work (e.g., Kong et al. [2024], Xia et al. [2022]), predominantly employ atlas-based paradigms that require learning diffeomorphic transformations from a predefined average template to target anatomies.

The field shows limited exploration of multi-modal approaches, with most studies focusing on single imaging modalities. Notable exceptions include works like Beetz et al. [2022b], which combines MRI and ECG data, and Biffi et al. [2019], which processed multiple cardiac views. This observation is particularly relevant given that 17 cardiac papers predominantly use MRI data, as seen in series of works by Beetz et al. [2022c], Beetz et al. [2022a], Beetz et al. [2024], while other anatomical structures often rely on different imaging modalities, such as CT scans for skull reconstruction [Sulakhe et al., 2022] and 3D scans for dental applications (Hosseinimanesh et al. [2023], Tian et al. [2022]). Most of the current methods emphasise static anatomical structure generation. Of all reviewed papers, only a small subset addresses dynamic anatomical modelling, with cardiac motion being the primary focus. Qiao et al. [2025, 2024] pioneered comprehensive approaches for generating dynamic cardiac shapes, incorporating temporal variations in their models. Their work demonstrates the feasibility of generating 3D+t anatomical models that capture both spatial and temporal variations in cardiac geometry.

The analysis reveals that topology handling varies significantly across anatomical structures. Vessel studies, though fewer in number, show the most diverse methodology usage, including diffusion-based approaches (Kuipers et al. [2024], Sinha and Hamarneh [2024]), reflecting the complexity of vascular topology. In contrast, cardiac studies, despite their larger number, show less methodological diversity in handling topological variations, with most works like Beetz et al. [2022a] and Seale et al. [2024] focusing on established VAE architectures.

Managing topology variations while maintaining anatomical validity represents a fundamental challenge. This is particularly evident in vessel generation tasks, where complex vessel features present unique challenges, as addressed in works like Feldman et al. [2023] and Wolterink et al. [2018]. Recent approaches using tree-structured architectures [Hu et al., 2021] and hierarchical frameworks [Hu et al., 2024a] to handle complex topological relationships.

The challenge of capturing fine-scale anatomical details remains significant, particularly for structures such as small vessels, valve features, classifications, and intricate surface geometries. Current methods often fail to preserve these details while maintaining global anatomical validity. Future research should explore multi-scale architectures specifically designed for detail preservation, coupled with novel loss functions that emphasise fine structural elements. Additionally, adaptive sampling strategies that allocate more computational resources to complex regions warrant investigation.

Ensuring the physiological plausibility of generated geometries requires addressing multiple constraints simultaneously, from local smoothness to global anatomical relationships. The field needs improved methods for incorporating physiological constraints into the generation process, validating the functional validity of generated geometries, and maintaining realistic relationships between different anatomical components.

### 7.2 Evaluations

The evaluation of deep learning-based virtual population generation models presents several significant challenges that require attention from the research community. These challenges can be categorised into three main areas: comprehensiveness of evaluation metrics, hierarchical assessment approaches, and clinical applicability.

First, current evaluation practices reveal an imbalanced and non-comprehensive approach to assessment. The geometric fidelity is well-addressed through point-wise and vertex-based measurements, but the topological characteristics often remain inadequately assessed. While there is a strong emphasis on fidelity metrics, other crucial aspects remain less thoroughly explored. Analysis of existing literature demonstrates this disproportionate focus on fidelity metrics, with multiple approaches developed for assessing geometric accuracy. These include comprehensive measurement approaches such as anthropometric analysis (Dou et al. [2025], Romero et al. [2021], Tian et al. [2022], Qiao et al. [2025], Wolterink et al. [2018], Qiao et al. [2024] and various similarity metrics. Statistical measures like COV [Zhang et al., 2024a, Sinha and Hamarneh, 2024] and MMD [Zhang et al., 2024a, Beetz et al., 2022b] provide quantitative assessments of distribution matching. However, the limited attention paid to utility, generalizability, and diversity metrics presents a significant gap in evaluating the practical value of generated populations. Moreover, the metrics for each aspect are yet to be standardised, making it difficult to understand and compare different models effectively.

Second, current evaluation approaches lack sufficient hierarchical depth in their assessment methodology. Many methods focus solely on either sample-level or population-level evaluation, failing to provide a comprehensive understanding of model performance across different scales. This gap is particularly evident in fidelity evaluation. For fidelity evaluation, most metrics are designed to compare the distributions of the real populations and the VPs, and specificity is the only sample-level metric. There is a demand to jointly use these two types of metrics for a hierarchical evaluation. From the utility side, the current approaches primarily rely on downstream tasks, including clinician validation [Abdi et al., 2019, Hwang et al., 2018], classification [Romero et al., 2021, Beetz et al., 2022b], and prediction [Huang et al., 2024]. While these downstream applications provide valuable insights, they often operate at a population level by the downstream models, missing the deeper implications of a single sample’s quality. This limitation is particularly critical for simulation-based applications where structural integrity and connectivity patterns determine the utility of generated populations.

Third, there is a notable gap in addressing the ISTs’ applicability and clinical outcome correlation of generated populations. While virtual population generation serves as a crucial foundation for ISTs, most current methods fail to demonstrate direct applicability to trial scenarios or establish clear connections to clinical outcomes. This disconnect manifests in several critical ways that require attention from the research community. There is also a scarcity of clinical validation in current evaluation approaches. Among the reviewed methods, only two papers Abdi et al. [2019], Hwang et al. [2018] have attempted to bridge this gap by incorporating clinical expertise into their evaluation process. These studies compared model-generated designs with clinician-created designs and sought expert validation of their results. The lack of widespread clinical validation raises questions about the practical utility of generated populations in realworld medical applications.

### 7.3 Future Directions

Based on the challenges discussed in the previous section, this section outlines key future directions for advancing virtual population generation methods. Our objective is to address each identified challenge systematically while providing actionable insights for researchers and practitioners in the field.

To better integrate multi-modal data for model training, multi-modal fusion frameworks from the field of computer vision and computer graphics can be adapted. Addressing multi-modal fusion is particularly crucial because the integration of complementary information across modalities could provide extra controls or constraints on the VP generation. Future research directions should focus on developing unified fusion architectures that can effectively integrate multi-modal data through cross-modal attention mechanisms. These architectures would address the current limitations in combining different types of anatomical information by establishing more sophisticated relationships between various modalities.

Specifically, three key modal combinations warrant investigation for VPs generation: 2D-3D visual fusion [Zhao et al., 2023], text-3D visual fusion [Yin et al., 2025, Zhao et al., 2023, Cheng et al., 2023], and text-2D visual fusion [Huang et al., 2023, Xia et al., 2021, Hu et al., 2024b]. These multi-modal fusion approaches could advance the field of anatomical geometry generation, leading to more accurate and controllable models. Beyond multimodal integration, two additional challenges need to be addressed: the lack of dynamic generation capabilities and the multi-resolution consistency maintenance of the generated meshes or point clouds. To address the first challenge, future research could adapt motion estimation models [Zhang et al., 2024b, Wang et al., 2023, Guo et al., 2022] to generate dynamic virtual populations, enabling the simulation of anatomical variations across different temporal states. For the second challenge, implementing shape completion [Zhao et al., 2021, Zhou et al., 2021, Wu et al., 2020] or 3D super-resolution [Pesavento et al., 2022, Li et al., 2021b, Hou et al., 2022] techniques could help maintain consistency across different resolution levels, ensuring accurate representation of anatomical structures at various scales. Beyond the specific methods discussed, broader advances in computer vision and shape generation could provide valuable new approaches for virtual population generation [Zeng et al., 2022, Shim et al., 2023, Metzer et al., 2023].

To address the challenge of complex topology handling, future research should focus on developing sophisticated topology-aware loss functions [Shit et al., 2021, Hu et al., 2024c, Jeong et al., 2023]. These functions would need to account explicitly for the topological characteristics of anatomical structures during the generation process. Additionally, implementing topology-adaptive models could provide a more flexible approach to handling various anatomical configurations [Gao et al., 2021, Yao et al., 2023, Bessadok et al., 2021, Banerjee et al., 2022].

The second challenge involves the generation of VPs under semantic control. The disentangled approach could associate specific meanings with specific latent variables [Eddahmani et al., 2023, Wang et al., 2024, Zhang et al., 2024c]. Therefore, it aligns well with human language and enables part-level edits based on intuitive concepts.

In the realm of anatomical geometry generation, preserving fine-scale anatomical details presents a significant challenge that requires innovative solutions. To address this challenge, attention-guided refinement emerges as a promising direction [Li et al., 2022, Zhang et al., 2023b, Tang et al., 2022]. This approach would leverage attention mechanisms to focus computational resources on regions requiring high-detail preservation. Beyond attention-guided methods, several additional approaches show promise [Xu et al., 2018, Yuan et al., 2023]. Multi-scale feature extraction could be employed to capture details at various levels of granularity [Díaz-Pernas et al., 2021, Saragadam et al., 2022, Wu et al., 2023].

In generating anatomically plausible VPs, incorporating both anatomical and anthropometric constraints is crucial for ensuring the generated shapes reflect realistic human characteristics. Anatomical constraints ensure that generated structures maintain proper medical and biological relationships. Anthropometric constraints ensure that generated anatomical structures conform to human body measurements and proportions.

The evaluation of the generated VP geometries presents significant challenges that require comprehensive and systematic approaches. Looking forward, the field needs to establish more rigorous and standardised evaluation methodologies that can effectively assess the quality and utility of generated anatomical structures at multiple scales. A key advancement in this direction would be the development of a systematic evaluation workflow that incorporates both sample-level and population-level metrics. This comprehensive framework should address four critical aspects: fidelity, utility, generalizability, and diversity. At the sample level, the evaluation would focus on individual geometric accuracy and anatomical correctness, while the population-level assessment would examine statistical distributions and variations across generated samples.

The evolution of evaluation methodology requires several key initiatives. First, comprehensive evaluation frameworks must be developed to balance all four aspects. This balanced approach would provide more complete assessments of generated populations and help researchers understand the trade-offs between these different aspects. The framework should include metrics that can quantify each aspect independently while also providing insights into their interactions. Second, the field needs to establish standardised metrics for both geometric and topological evaluation. These metrics would enable meaningful comparisons between different generation approaches and facilitate reproducible research. Third, simulation-based validation protocols need to be integrated into the evaluation process. These protocols would strengthen the connection between evaluation metrics and practical applications, ensuring that generated anatomical structures function as intended in simulated environments for ISTs.

To enhance clinical translation, the field should implement a continuous improvement loop consisting of virtual population generation, systematic evaluation, and ISTs validation. This iterative process would ensure that generated anatomical structures not only meet technical quality metrics but also satisfy practical clinical requirements. The feedback from in silico trials would inform improvements in both generation methods and evaluation criteria, leading to progressively more reliable and clinically relevant virtual populations.

## 8 Conclusion

This review systematically categorised deep learning methods for generative modelling of virtual patient populations into six primary approaches: VAE-based, GAN-based, flow-based, diffusion-based, DeepSSM-based methods, and other approaches. We established comprehensive taxonomies for both generation strategies (unconditional versus conditional) and evaluation frameworks encompassing fidelity, utility, generalisability, and diversity metrics.

Our analysis reveals significant methodological imbalances across anatomical domains. Whilst cardiac applications demonstrate sophisticated methodologies, complex vascular structures with diverse topologies remain underexplored. Similarly, evaluation practices disproportionately emphasise geometric fidelity whilst neglecting utility, generalisability, and diversity assessments. The lack of standardised evaluation metrics across studies further constrains meaningful performance comparisons.

Key architectural limitations emerged from our cross-anatomical analysis. VAE-based methods, though effective for fixed-topology cardiac structures, struggle with complex vascular geometries characterised by variable branching patterns. GAN-based approaches excel at preserving fine details but face challenges in controlled generation of anatomically coherent structures. Additional limitations include insufficient multi-modal integration, inadequate handling of dynamic anatomical variations, and persistent difficulties balancing detail preservation with global anatomical validity.

Three critical evaluation challenges impede comprehensive assessment: disproportionately focus on geometric fidelity metrics, absence of standardised measures across assessment dimensions, and limited hierarchical evaluation spanning sample-to-population scales. Most critically, current methods rarely demonstrate clinical applicability through ISTs or establish clear connections to clinical outcomes.

Based on these findings, we identify three crucial research directions: developing methods capable of handling complex anatomical structures with diverse topologies, establishing standardised evaluation protocols addressing all four assessment dimensions, and bridging the translation gap between virtual population generation and clinical applications through robust simulation-based validation frameworks.

The convergence of advanced generative modelling with virtual population synthesis represents a transformative opportunity for personalised medicine and in silico clinical trials. As the field matures beyond proof-of-concept demonstrations towards clinical translation, the systematic frameworks and research priorities outlined in this review provide a roadmap for realising the full potential of virtual patient populations in accelerating medical innovation whilst reducing reliance on traditional clinical trial paradigms.

## Supporting information

Supplementary Table S1. Data Extraction Sheet

Supplementary Figure S1. PRISMA 2020 Flow Diagram

Supplementary File S1. Search Strategies

Supplementary File S2. Data Extraction Explanation

Supplementary File S3. Inclusion Exclusion Criteria

## Data Availability

All data produced in this work are contained within the manuscript and its supplementary materials (extraction tables, search strategies, PRISMA flow, and exclusion lists).
This study did not generate new human participant data. Quantitative results were extracted from previously published articles; the curated extraction tables and meta-analysis inputs are provided in the manuscript

## 9 Acknowledgements

AFF acknowledges support from the Royal Academy of Engineering under the RAEng Chair in Emerging Technologies (INSILEX CiET1919/19), ERC Advanced Grant – UKRI Frontier Research Guarantee (INSILICO EP/Y030494/1), the UK Centre of Excellence on insilico Regulatory Science and Innovation (UK CEiRSI) (10139527), the National Institute for Health and Care Research (NIHR) Manchester Biomedical Research Centre (BRC) (NIHR203308), the BHF Manchester Centre of Research Excellence (RE/24/130017), and the CRUK RadNet Manchester (C1994/A28701).

AFF is co-founder, shareholder and board member of adsilico Ltd and OculomeX Health Ltd. Other authors have no conflicts to disclose.

## Footnotes

1 www.prisma-statement.org

