## Supplementary figures and images for "Synthetic Anatomy: Deep Learning Models for Virtual Population Generation–A Review"

### Supplementary Figure S1. PRISMA 2020 Flow Diagram

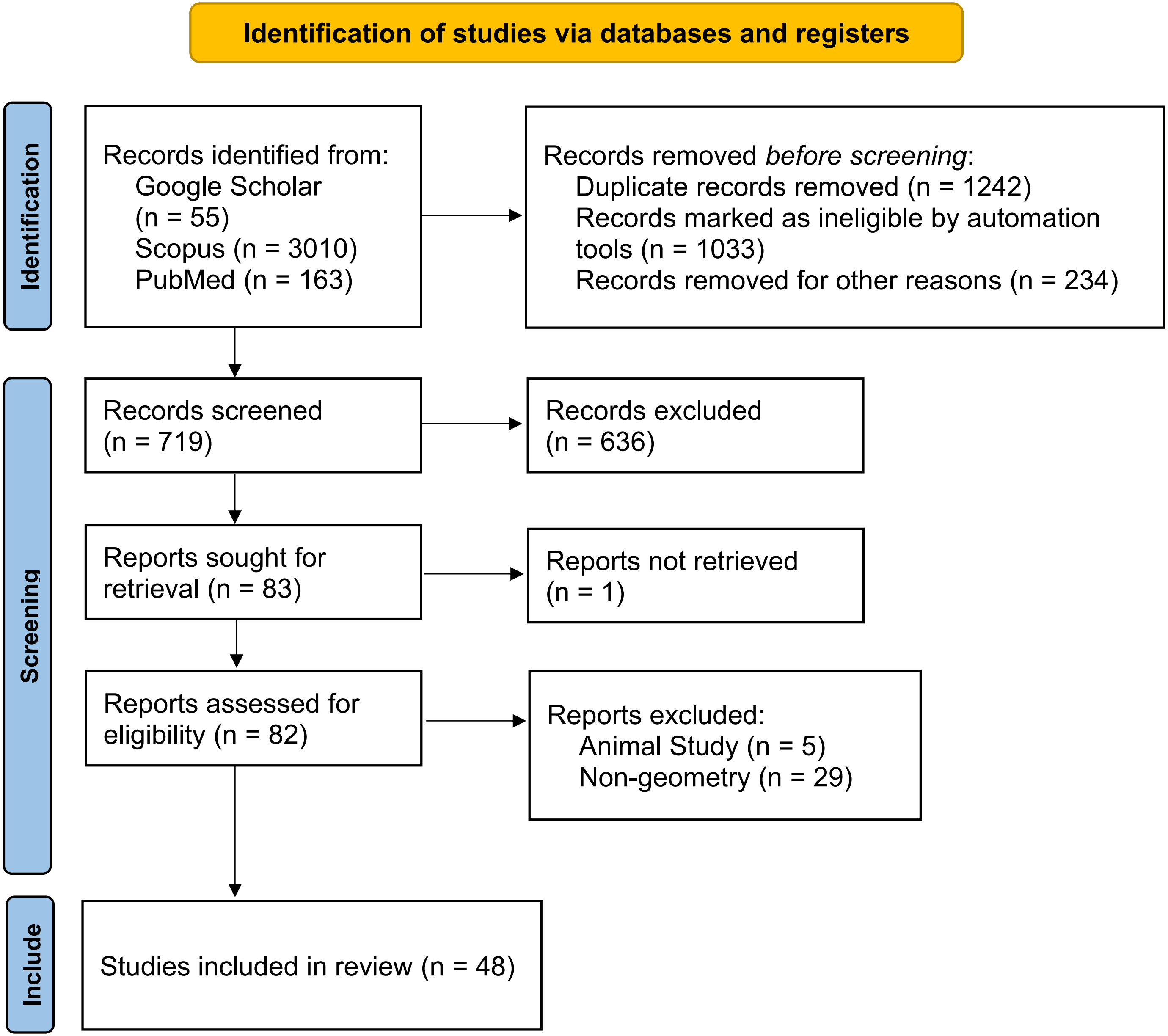
