## Supplementary File S1. Search Strategies for "Synthetic Anatomy: Deep Learning Models for Virtual Population Generation–A Review"

*Search Strategy：*

We conducted a systematic search in PubMed, Scopus, and Google Scholar from inception to [2015-06-01]. English language restrictions were applied. The PRISMA flow diagram is reported in Supplementary Figure S1. PRISMA 2020 Flow Diagram (Figure 4 in the manuscript)

*Year*: 2015-2025

*Title, abstract, keywords*: (GAN OR VAE OR diffusion OR normali* flow OR generative) AND (4D OR 3D OR Shape OR geometry OR SDF OR implicit) AND (clinical OR anatom* OR cardi* OR vessel OR vascular OR bone OR lung OR jaws OR dental OR teeth OR brain OR ribs)

Scopus:3010 (article, conference paper, book chapter, book, final version, English, using the keyword above)

PubMed:163 (book and documents, using the keywords above)

Google Scholar: 55 (generative models clinical, OR anatomic, OR cardiovascular, OR heart, OR vessel, OR bone, OR lung, OR jaw, OR dental, OR teeth, OR rib, OR brain)
