## Supplementary File S2. Data Extraction Explanation for "Synthetic Anatomy: Deep Learning Models for Virtual Population Generation–A Review"

*Data extraction & items*

We developed a piloted extraction form capturing study metadata, anatomy, model family (GAN/VAE/diffusion/GNN), topology handling, multimodal inputs and outputs, training datasets size (Table 1 to Table 11 in the manuscript), and metrics mapped to four dimensions—fidelity, utility, generalisability, diversity (Table 12 to Table 15 in the manuscript). We directly adopted the experimental results from the paper included in our review and performed a rough meta-analysis (Table 16 to Table 19 in the manuscript). For your convenience, we summarised Table 1 to Table 11 in Supplementary Table S1. Data Extraction Sheet which contains the 48 studies included in our review.
